# An integrated multi-omics investigation of W-NK1, a cytokine-primed non-engineered natural killer cell therapy product

**DOI:** 10.1101/2024.07.08.24310018

**Authors:** Laura Arthur, Nitin Mahajan, Jayakumar Vadakekolathu, Tom Leedom, David J. Boocock, Clare Coveney, Alex Hamil, Kristann Magee, John Dean, Elizabeth Schramm, Benjamin Capoccia, Vincent Petit, Nupur Bhatnagar, Christian Pinset, Awais Younis, Craig Doig, Benjamin Thomas, Evangelia Williams, Lena Luukkonen, Yanira Ruiz-Hereida, Alejandro Martin Munoz, Paula Comune Pennacchi, Daniel Primo, Neysa Dagostino, Stacy K. Lewis, Natasha Edwin, John Muth, Melissa Berrien-Elliott, Todd A. Fehniger, Jan K. Davidson-Moncada, Sergio Rutella

## Abstract

**Background:** Natural killer (NK) cells originate from bone marrow precursors and mediate effective anti-tumor responses. Clinical trials of cytokine-primed memory-like (ML) NK cells in acute myeloid leukemia (AML) have demonstrated activity without major toxicity, including graft-versus-host disease or cytokine release syndrome. However, broad application of non-expanded, non-engineered ML NK cells has been hindered by limited availability of NK cells from a single donor, thereby precluding aggressive dose escalation and repeat dosing. W-NK1 is derived from human peripheral blood mononuclear cells undergoing ML reprogramming with a proprietary heteromeric fusion protein complex including IL-12, IL-15 and IL-18.

**Methods:** We conducted a multi-omics characterization of W-NK1 by interrogating its transcriptomic, proteomic and metabolic profile. Using functional assays, we assessed W-NK1’s cytotoxicity under adverse culture conditions, as well as W-NK1’s trafficking and killing abilities in immunodeficient mice engrafted with THP-1 AML. Finally, we evaluated W-NK1’s phenotype and in vivo expansion kinetics in one patient with AML enrolled in study NCT05470140.

**Results:** W-NK1 displayed an activated, hyper-metabolic, and proliferative state differing from unstimulated conventional NK cells (cNK) from healthy donors. When compared to external single-cell NK datasets, W-NK1 was largely annotated as NKG2A^+^ and showed low relatedness with adaptive NK states characterized by HCMV-induced inflammatory memory. W-NK1 outperformed cNK cells in terms of in vitro killing of a broad panel of AML cell lines, with no appreciable cytotoxicity against normal cell lines. The expression of nutrient transporters was higher in W-NK1 compared to cNK cells and was retained even in adverse culture conditions designed to mimic an immunosuppressive tumor microenvironment. In mice engrafted with THP-1 AML, W-NK1 trafficked and efficiently homed to the bone marrow, where it mediated better tumor control than cNK cells. W-NK1 expanded, underwent phenotypic changes and persisted with effective elimination of circulating AML blasts through day 14 after infusion in one patient treated on clinical trial NCT05470140. Immunofluorescence staining of BM sections collected on day 28 showed increased expression of both CD56 and CD3 compared to a pre-treatment biopsy.

**Conclusions:** Our study offers a comprehensive characterization of W-NK1 as an effective cell therapy product for AML and solid tumor malignancies.

**What is already known on this topic:** Natural killer (NK) cells have been shown to be safe and effective for treating certain human malignancies. Nonetheless, limitations for adoptive cell therapy exist which include trafficking / homing to tumor tissues as well as metabolic resilience in an adverse microenvironment.

**What this study adds:** W-NK1 is distinct transcriptionally and functionally from conventional NK cells with improved anti-tumor effector functions and metabolic adaptation in hostile culture conditions. Moreover, W-NK1 was readily detectable post-infusion in a patient with refractory acute myeloid leukemia.

**How this study might affect research, practice or policy:** Our in vitro and in vivo findings indicate that W-NK1 is an effective NK-cell therapy product and augur positively for patients being treated in phase I immunotherapy clinical trials.

## INTRODUCTION

Natural killer (NK) cells are effector innate lymphoid cells originating from bone marrow (BM) progenitors and protect the host from infection as well as mediating anti-tumor responses. Conventional NK (cNK) cells circulate in blood, traffic through most tissues and can be rapidly mobilized to sites of disease. A primary function of NK cells is to directly recognize diseased cells through activating NK cell receptors that detect cellular stress ligands or the loss of self-identifying major histocompatibility complex (MHC) class I proteins.^1^

Human NK cells are usually classified into two main subsets, i.e., CD56^bright^CD16^-^ cells which produce abundant cytokines and become cytotoxic upon IL-15 priming, and CD56^dim^CD16^+^ cells which are cytotoxic at rest and mediate antibody-dependent cellular cytotoxicity (ADCC).^2^ Single-cell RNA-sequencing (sc-RNA-seq) approaches are enabling a more granular classification of blood and tissue-resident NK-cell populations, including type 1 NK (NK1) cells, corresponding to CD56^dim^CD16^+^ cells, NK2 cells resembling CD56^bright^CD16^-^ cells, and NK3 cells comprising CD16^dim^ adaptive NKG2C^high^ NK cells with memory-like (ML) properties and enhanced functional responses to a variety of stimuli, including bacteria, viruses, cytokines and haptens.^1^ NK cells can directly kill tumor targets through the perforin/granzyme pathway or induce apoptosis by ligating death receptors such as Fas-ligand, TNF or TRAIL. Upon activation, NK cells proliferate and produce cytokines such as interferon (IFN)-γ and tumor necrosis factor (TNF)-α, as well as chemokines such as CCL3, CCL4, CXCL8, and CCL5, which are crucial for orchestrating adaptive immune responses. Clinical reports have indicated that NK cell products are safe,^3–6^ potentially overcoming the limitations of T cell-based cellular immunotherapies. In light of their unique functional attributes, NK are being harnessed for cancer treatment, as they can be readily isolated in large quantities from donor leukapheresis products and produced off-the-shelf for clinical validation.^7^

It has now evident that NK cells exhibit various forms of innate memory, contingent upon the initial NK population and stimulation conditions. Three types of NK-cell memory have been reported: antigen-specific, cytokine-induced, and liver-restricted. Clinically, cytokine-induced ML NK cells exhibit several advantages over cNK-cell platforms. Human ML NK cells that differentiate after IL-12, IL-15, and IL-18 stimulation exhibit enhanced cytokine production, cytotoxicity, proliferation and persistence, as well as the ability to eliminate tumor cells *in vivo* without the need for additional engineering.^8–10^ Clinical trials of ML NK cells in acute myeloid leukemia (AML) reported no graft versus host disease (GvHD), cytokine release syndrome (CRS), immune effector cell-associated neurotoxicity syndrome (ICANS), or severe treatment related adverse events (Grade ≥3),^9^ indicating that the enhanced functionality imparted on ML NK cells by cytokine priming does not compromise the safety profile typically reported with other non-engineered NK-cell platforms.^3,4^ However, the use of ML NK cells as a point-of-care, non-expanded NK therapy has been limited to one donor providing a cellular therapy to one patient, thereby restricting the potential for aggressive dose escalation, repeat dosing, and broader therapeutic application.^11^

W-NK1 is a non-engineered NK cell derived from human peripheral blood mononuclear cells (PBMCs) undergoing ML reprogramming through cytokine stimulation with a proprietary heteromeric fusion protein complex, including IL-12, IL-15 and IL-18 recombinant proteins,^11^ followed by expansion in a feeder-cell-free culture system.^9^ W-NK1 is cryopreserved to yield an off-the-shelf allogeneic ML NK product with anti-tumor cell activity. Due to controlled *ex vivo* cellular expansion, W-NK1 can treat multiple patients with repeated doses from a single apheresis product, thereby enhancing patient access to ML NK therapy.

In this study, we conduct a multi-omics characterization of W-NK1 by interrogating its transcriptomic, phenotypic, and metabolic profile. Using functional assays, we assess the *in vivo* and *in vitro* cytotoxicity of W-NK1 under adverse culture conditions, including tumor microenvironment (TME)-aligned culture media, patient-derived malignant ascites, and hypoxia. Our findings suggest that W-NK1 might address previously reported challenges in adoptive cell therapy^12^ and lay the groundwork for positioning W-NK1 as an effective non-engineered NK-cell product for treating human malignancy.

## RESULTS

### W-NK1 acquires a unique phenotype and exhibits high activation and proliferation

We initially utilized spectral flow cytometry to characterize W-NK1’s phenotype, including the expression of chemokine receptors, activating/inhibitory receptors and NK-cell maturation and effector function molecules (figure 1A and table S1). When compared to unstimulated NK cells (herein referred to as cNK), W-NK1 exhibited a relatively immature phenotype with heightened expression of CD56 and reduced expression of CD16, CD45RA, CD57, and NKp80 (figure 1B).^13,14^ Furthermore, CD69 and CD25 were markedly elevated in W-NK1, aligning with the previously noted phenotype of cytokine-stimulated ML NK cells.^9^ The proliferative nature of W-NK1 was exemplified by the enhanced expression of Ki67, and its cytolytic capacity was supported by the expression of perforin and GZMB. Furthermore, activating receptors NKG2D, CD244, and CD226, as well as the co-activating receptor CD2, were significantly up-regulated on W-NK1 compared to cNK from the same donors. Highlighting their distinction from human cytomegalovirus (HCMV)-adaptive NK cells, W-NK1 showed minimal expression of NKG2C. NK inhibitory receptors, which attenuate effector responses when binding their ligands, displayed a complex pattern on W-NK1, with KLRG1 and CD328 (Siglec-7) being down-regulated and TIM3, CD96 (TACTILE), and NKG2A being significantly over-expressed in W-NK1 compared with cNK. Moreover, W-NK1 showed significantly increased expression of cell adhesion molecules CD11A and CD11C, as well as the chemokine receptor CCR2. Importantly, W-NK1 also exhibited increased expression of CD86, CD100, and NKp30, which are known to engage T cells, B cells, and dendritic cells, respectively (figure 1B).^15,16^ Given the role for chemokine receptors in cell trafficking to tumor sites,^17^ we conducted a more in-depth assessment of their expression by flow cytometry. The frequency of CXCR4, CXCR3, and CCR7 positive cells was significantly higher in W-NK1 compared to cNK (n = 3; figure 1C). Taken together, the phenotype of W-NK1 supports its functional characterization as a highly activated and cytotoxic NK cell, coupled with the potential for enhanced trafficking and immune system engagement.

**Figure 1.**
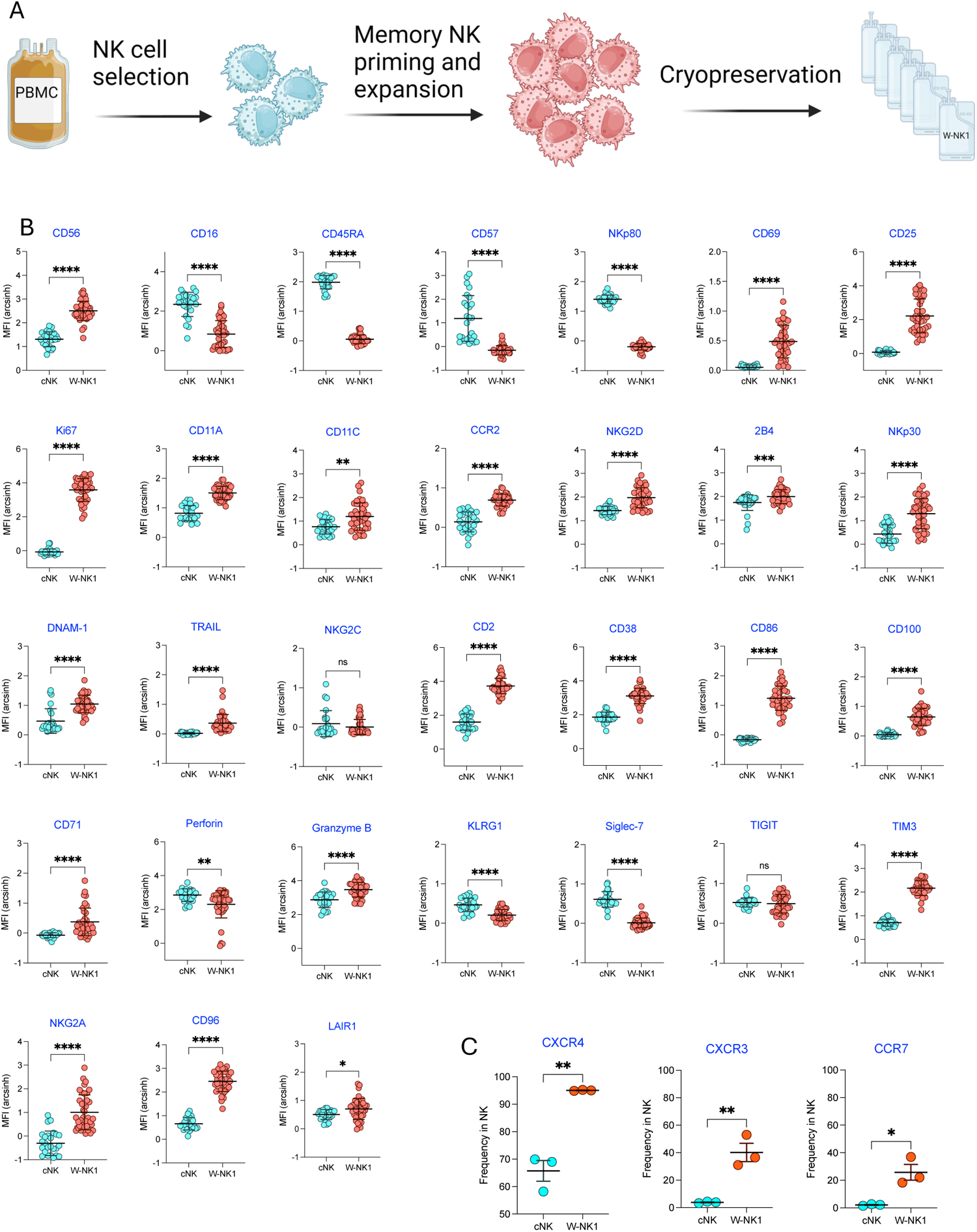
W-NK1 phenotype by spectral flow cytometry. (A) W-NK1 manufacturing schema. (B) Median fluorescence intensity (MFI) of selected surface and intracellular markers expressed by cNK and W-NK1 generated from the same donors. Expression intensity was arcsin-transformed prior to analysis. Bars represent the mean ± standard error of the mean (SEM). Mean MFIs were compared using an unpaired t-test. (C) Frequency of cNK and W-NK1 expressing the indicated chemokine receptor (CXCR4, CXCR3, and CCR7) as assessed by flow cytometry in three paired NK samples. Bars represent the mean ± SEM. Mean frequencies were compared by a paired t-test.

### W-NK1’s transcriptional rewiring supports metabolic fitness

We next performed bulk RNA-sequencing to discern transcriptomic disparities between W-NK1 and cNK cells. Following extensive QC and filtering, we identified a set of 7,061 differentially expressed genes (DEGs) at a false discovery rate (FDR) < 0.01. Among the genes upregulated in W-NK1 were *IL2RA*, *TPX2* and *MKI67*, indicating the establishment of an activated and proliferative state after cytokine expansion, as well as molecules involved in NK-cell regulation, such as *CSF2* encoding GM-CSF, and *BIRC5* encoding survivin (figure 2A).

**Figure 2.**
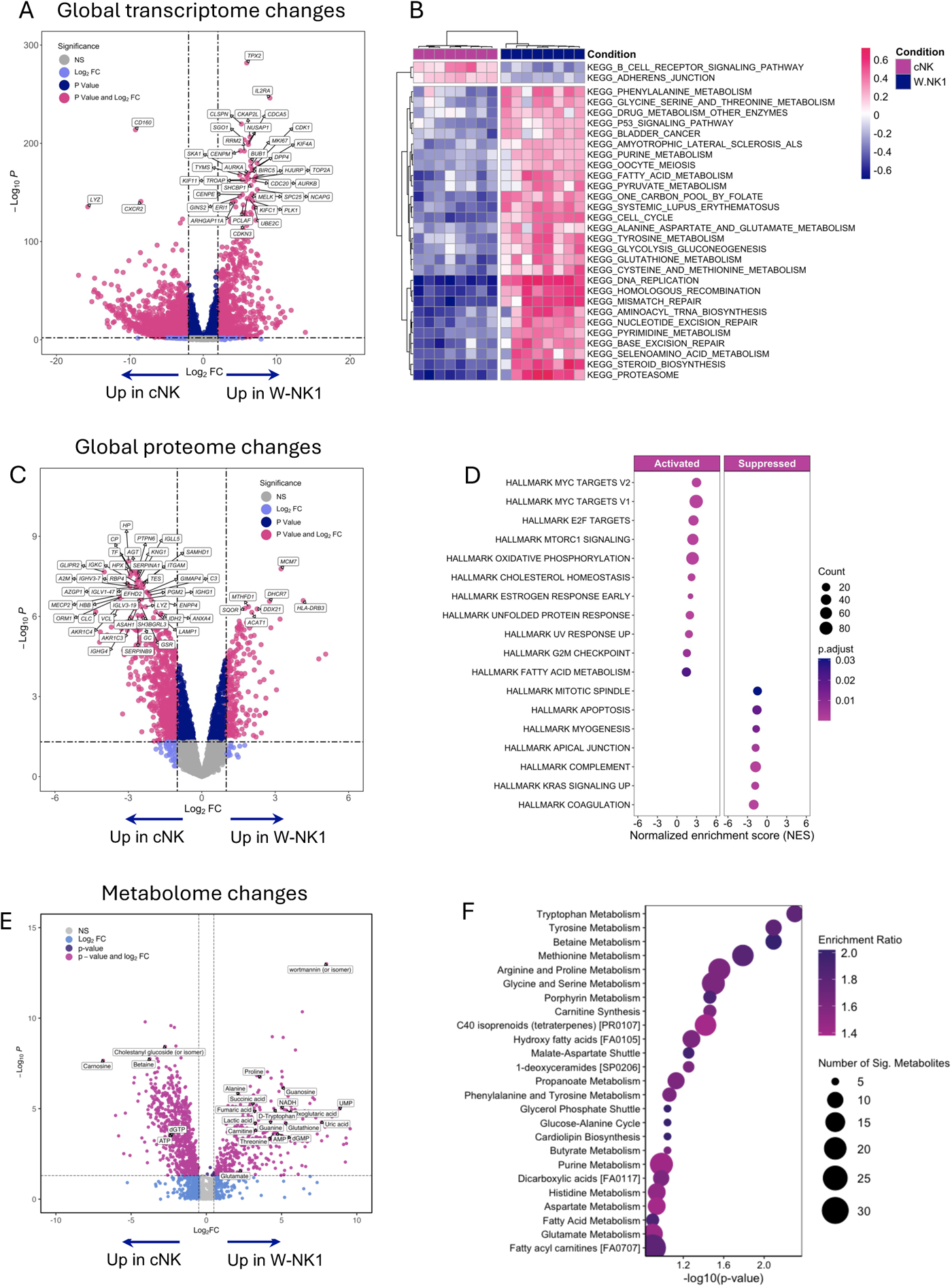
Multi-omics characterization of W-NK1. (A) Eight matched cNK and W-NK1 products were used for bulk RNA-sequencing. Volcano plot displaying the DEGs between cNK (left) and W-NK1 (right) at an FDR < 0.01. The top 40 DEGs are highlighted, with each purple dot representing an individual gene exhibiting an adjusted *P* < 0.01 and a log2-fold change >2.0. (B) Heatmap showing the differentially expressed KEGG pathways from GSVA in cNK and W-NK1 (FDR < 0.01). (C) Volcano plot presenting the DE proteins in lysates of cNK (left) and W-NK1 (right) at FDR < 0.01. The top 40 DE proteins are highlighted, with each purple dot denoting an individual protein exhibiting an adjusted *P* < 0.05 and a log2-fold change >1.0. (D) Dot plot summarizing the normalized enrichment scores (NES) of the 50 hallmark pathways from the MSigDB (clusterProfiler package in R). DE proteins between cNK and W-NK1 were used as input. (E) Volcano plot presenting DE metabolites in lysates of cNK (left) and W-NK1 (right) at FDR <0.01 and log2-fold change >2.0. (F) Dot plot of the enrichment ratio and number of significantly differentially expressed metabolites in donor-matched W-NK1 and cNK (n = 10).

Gene set variation analyses (GSVA) revealed that, compared to cNK cells, W-NK1 was significantly enriched in glycolysis, amino acid, and fatty acid metabolism KEGG pathways, as well as DNA replication and cell cycle-related gene programs (figure 2B). This observation implies the potential for W-NK1 to surmount metabolic constraints imposed by the hostile AML TME.^18^ The above transcriptomic findings were largely supported by mass spectrometry-based quantification of relative protein abundance in cell lysates. This analysis identified 1,598 differentially expressed proteins between W-NK1 and cNK (figure 2C), as well as enrichment in oxidative phosphorylation, lipid homeostasis and cell cycle progression Hallmark pathways. The latter included *MYC* signaling (figure 2D), which promotes NK metabolic reprogramming and mediates NK functional responses in mice.^19^

When assessing relative metabolite abundance in paired W-NK1 and cNK lysates by mass spectrometry, we observed that W-NK1’s bioenergetic pathways are shifted towards amino acid biosynthesis and lipid metabolism (figure 2E), with tryptophan, tyrosine, and methionine metabolism being highly enriched in W-NK1 (figure 2F). These adaptations may support high rates of glycolysis in W-NK1 or contribute essential substrates for its proliferation and expansion, analogous to the “Warburg metabolism” described in cancer cells.^20^

### W-NK1 is transcriptionally heterogeneous

We next explored the cellular heterogeneity of W-NK1 with single-cell resolution (figure S1A). Transcriptome analyses revealed decreased intra-donor variance of cluster proportions in W-NK1 compared to cNK cells, as well as tighter clustering of W-NK1 in a primary component analysis (figure S1B-1D). Overall, W-NK1 exhibited higher expression of *NCAM1, MKI67, ILR2A* and low expression of *FCGR3A* (figure S1E), in agreement with bulk RNA-seq and spectral flow cytometry data. Unbiased clustering and Uniform Manifold Approximation and Projection (UMAP) visualization of scRNA-seq data performed on 49,203 high-quality cells (25,374 W-NK1 and 23,829 cNK) from four healthy donors resulted in eight cellular states (c0-c7; figure 3A-B). The top 5 markers selected for cluster annotation are shown in figure S2A-B and listed in table S3. Cluster 0 comprised the predominant population, representing 38.7% of total NK cells. These cells demonstrated increased expression of key cytotoxicity genes, including *FGFBP2*, *KLRF1*, *GZMM* and *SPON2* (table S3), thereby supporting and extending previous high-dimensional analyses of NK populations^21^ as well as recapitulating the transcriptional features of the recently described NK1 subset of human blood NK cells.^22^ To identify differentially expressed genes between W-NK1 and cNK cells in cluster 0, whilst mitigating the risk of false discoveries and accounting for the intrinsic variability of biological replicates,^23^ we generated pseudobulk expression data from the sc-RNA-seq runs. This analysis revealed 6,567 over-expressed genes at an FDR <1% in ‘cytotoxic’ c0 cells from W-NK1 compared to their cNK counterpart (figure S2C). These genes were enriched in biological processed related to cell cycle progression, pro-inflammatory cytokine signaling and cellular metabolism (figure S2D).

**Figure 3.**
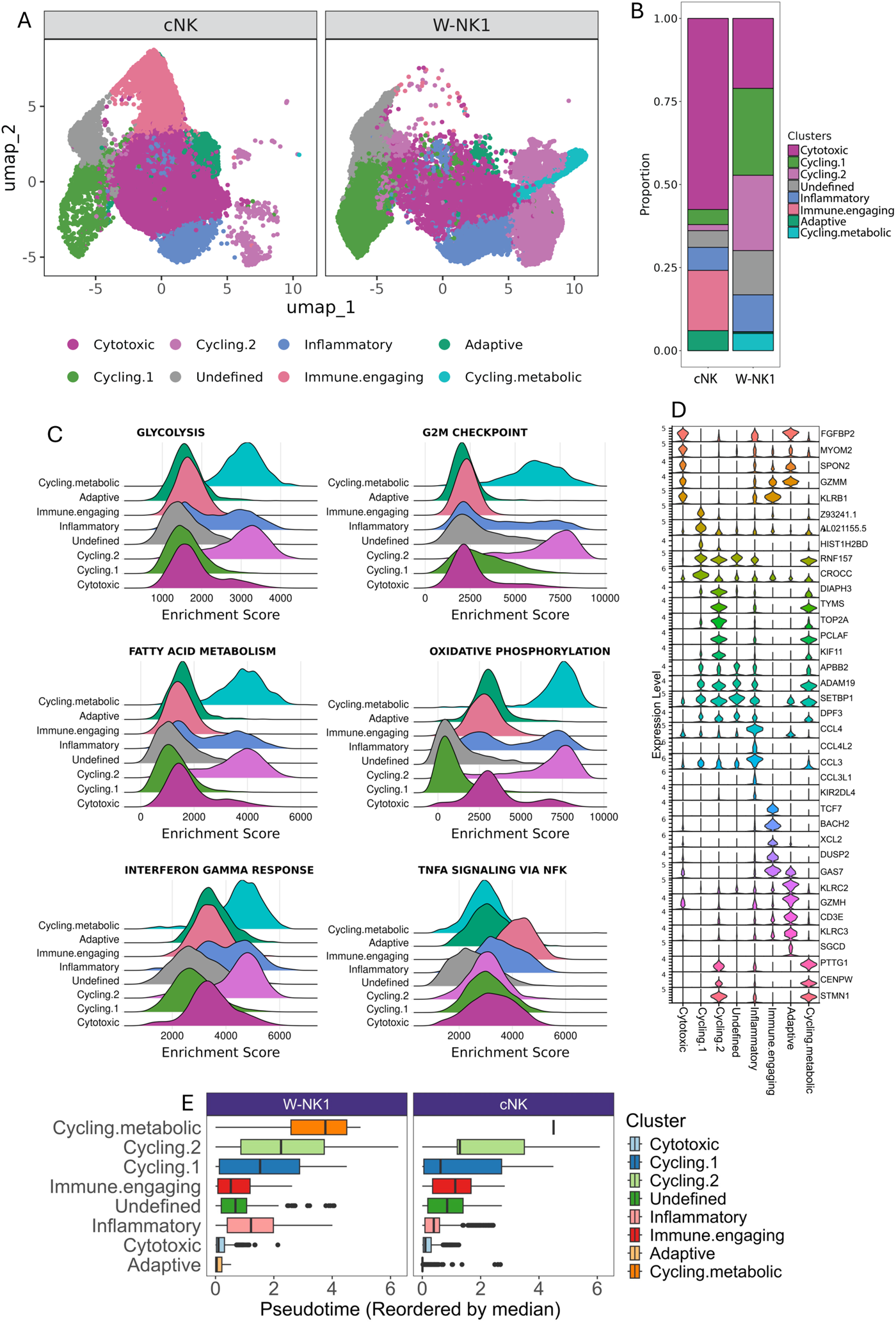
Single-cell transcriptomic analysis of W-NK1. (A) Uniform Manifold Approximation and Projection (UMAP) visualization of 49,203 high-quality NK cells (23,829 cNK and 25,374 W-NK1), highlighting 8 transcriptionally distinct cell states. (B) Linear modeling scRNA-seq data employing the *propeller* function in speckle to estimate differences in cell cluster proportions between W-NK1 and cNK cells. (C) Ridge plots showing metabolism-related and cell cycle-related Hallmark pathway enrichment scores, which were calculated using the escape package in R. (D) Violin plots depicting the expression of selected markers in W-NK1 and cNK. (E) Bar graphs of pseudotime values in W-NK1 and cNK cells. Transcriptional trajectory analyses were conducted using the Monocle3 package in R. Clusters were ordered based on the inferred pseudotime.

We then employed linear modeling of scRNA-seq data (*propeller* function in speckle) to estimate differences in cell cluster proportions between W-NK1 and cNK cells.^24^ As summarized in figure 3B and table S3, all clusters except c4 were significantly differentially abundant in W-NK1 compared to cNK cells. Cluster c7 was exclusively detected in W-NK1 and comprised dividing and metabolically active cells expressing *HMGB3, CDKN3, CCNB1, CCNB2,* and *ENO1* as well as glycolysis and oxidative phosphorylation gene programs (figure 3C-D). Genes associated with DNA replication and repair, as well as cell cycle control, including *STMN1, TYMS, PCLAF,* and *TK1*, were up-regulated in c7 and c2, with the latter also showing high expression of genes mediating cytoskeletal modeling, such as *TOP2A, TPX2,* and *CDK1*. Cluster 1 highly expressed the proliferation marker *MKI67*, along with several histone-coding genes and non-coding RNAs (table S3). Cells in c3 showed a *CD56^high^, CD16^low^* and *GZMB^+^* transcriptional profile and expressed metalloproteinases (*ADAM19*) and ring finger protein *(RNF157*), in conjunction to genes associated with DNA replication (*APBB2, DPF3,* and *SETBP1*; figure 3D). Whereas c4 was enriched in chemokine and inflammatory cytokine-encoding genes such as *CCL4, CCL4L2, CCL3,* and *IFNG* (figure 3D), c5 cells exhibited expression of *GZMK*, *TCF7* and dendritic cell-attracting chemokines *XCL1* and *XCL2*, which might promote NK-cell survival by modulating *GZMB* expression (18, 19).

We then constructed a single-cell trajectory using Monocle3. Pseudotime inference outlined a clear progression of maturation stages from cytotoxic W-NK1 in c0 to ‘hyper-metabolic’ and proliferative transcriptional states in c8, as a result of cytokine priming (figure 3E). In aggregate, single-cell profiling suggested that W-NK1’s activated state, as well as its enhanced metabolic flexibility, could underpin persistence and potent anti-tumor effector functions in a nutrient-deprived TME.

### Comparison of W-NK1 to external datasets using transfer learning

To unbiasedly define the population structure of W-NK1, we first projected sc-RNA seq data onto a previously constructed transcriptional map of 49,530 blood NK cells from HCMV-positive and HCMV-negative healthy donors (generated using the same chemistry and technology as in our NK dataset; GSE197037).^25^ The NK map includes NKG2C^+^ adaptive NK cells which expand in HCMV^+^ individuals and after *in vitro* imprinting by HCMV peptides and pro-inflammatory cytokines (figure S3A). As shown in figure S3B, W-NK1 was enriched in proliferation-related gene programs and showed depletion of both CD56^dim^ and CD56^bright^ NK clusters, with no detectable change in adaptive gene signatures (figure S3B). This observation suggests low relatedness between W-NK1 and adaptive NK-cell states characterized by HCMV-induced inflammatory memory. Label transfer of W-NK1 cells onto the HCMV dataset using an orthogonal computational approach^26^ corroborated these findings by revealing a greater than 20.0-fold expansion of proliferative functional states in W-NK1 compared to cNK (figure S3B) as well as identifying 128 DEGs in W-NK1 relative to the HCMV map. This gene set included molecules involved in IFN-γ signaling along with pro-inflammatory chemokines such as *CCL3* (figure S3C).

We then projected NK cells onto a novel reference map of 44,640 healthy donor-derived NK cells (https://zenodo.org/doi/10.5281/zenodo.8434223)^27^ and we found that W-NK1 was largely annotated as NKG2A^+^, in stark contrast to cNK cells (figure S3D). These features recapitulate previous reports of cytokine-induced ML NK cells, where NKG2A was identified as a dominant, transcriptionally induced checkpoint endowed with crucial roles in NK-cell responses to cancer.^9^ In aggregate, these data point to unique transcriptional features of W-NK1 which set it apart from previously developed cell therapy products.

### W-NK1 potently and selectivity eliminates malignant cells in vitro

To evaluate W-NK1’s function against AML, we conducted *in vitro* cytotoxicity assays using HL-60, THP-1, and TF-1 cells, which originate from patients with acute promyelocytic leukemia, acute monocytic leukemia, and acute erythroid leukemia, respectively. Compared to cNK cells, W-NK1 showed a significantly greater killing ability when co-cultured with HL-60 AML cells for 2 days over a broad range of effector-to-target (E:T) ratios (figure S4A). Cytotoxicity assays of paired W-NK1 and cNK generated from 5 independent donors revealed that W-NK1 is ∼3-fold more potent (mEC50 1.7 and 5.2; *P = 0.032*; figure S4A). Efficient cytotoxicity was also demonstrated against THP-1 and TF-1 AML cells in a 72-hour co-culture assay, although variable levels of sensitivity were observed across cell lines (figure S4B). While THP-1 was highly sensitive and showed dose-dependent W-NK1 cell-mediated killing, extensive killing of TF-1 cells by W-NK1 was only documented at the highest E:T ratio. We next assessed cytotoxic activity against several solid tumor cell lines, including head and neck squamous cell carcinoma (SCC-25), colorectal (LoVo), and ovarian cancer (SKOV3). These experiments showed that W-NK1 efficiently kills solid tumor targets, as indicated by the significantly decreased number of viable tumor cells remaining in the cultures after 24 hours at a 3:1 E:T ratio, as compared to ‘no-effector’ control conditions (figure S4C). Importantly, W-NK1 did not exert any cytotoxicity against non-transformed cell types (figure S4D). In aggregate, cytotoxic assays revealed that W-NK1 efficiently lyses tumor samples while retaining the ability to discern between malignant and benign target cells and suggest its potential for decreased off-tumor activity and improved safety profile.

### W-NK1 retains high cytotoxicity under culture conditions mimicking metabolically hostile tumor microenvironments

Several TME features hinder immune cell function, including nutrient-deprivation/scarcity, hypoxia, and the abundance of immunosuppressive agents.^18,28,29^ We set out to evaluate the potential impact of these factors on W-NK1’s function by assessing its cytotoxicity under metabolically hostile conditions. To this end, we utilized a ‘TME-aligned’ culture medium, which was obtained by stimulating human mesenchymal stromal cells (MSCs) with TNF-α^30^ and is characterized by an acidic (pH = 6.9) and hypoglycemic milieu (6.0 mM glucose), as well as high concentrations of immunosuppressive mediators, such as nitric oxide, PGE_2_, IDO, IL-10, and TGF-ϕ31 (figure 4A).

**Figure 4.**
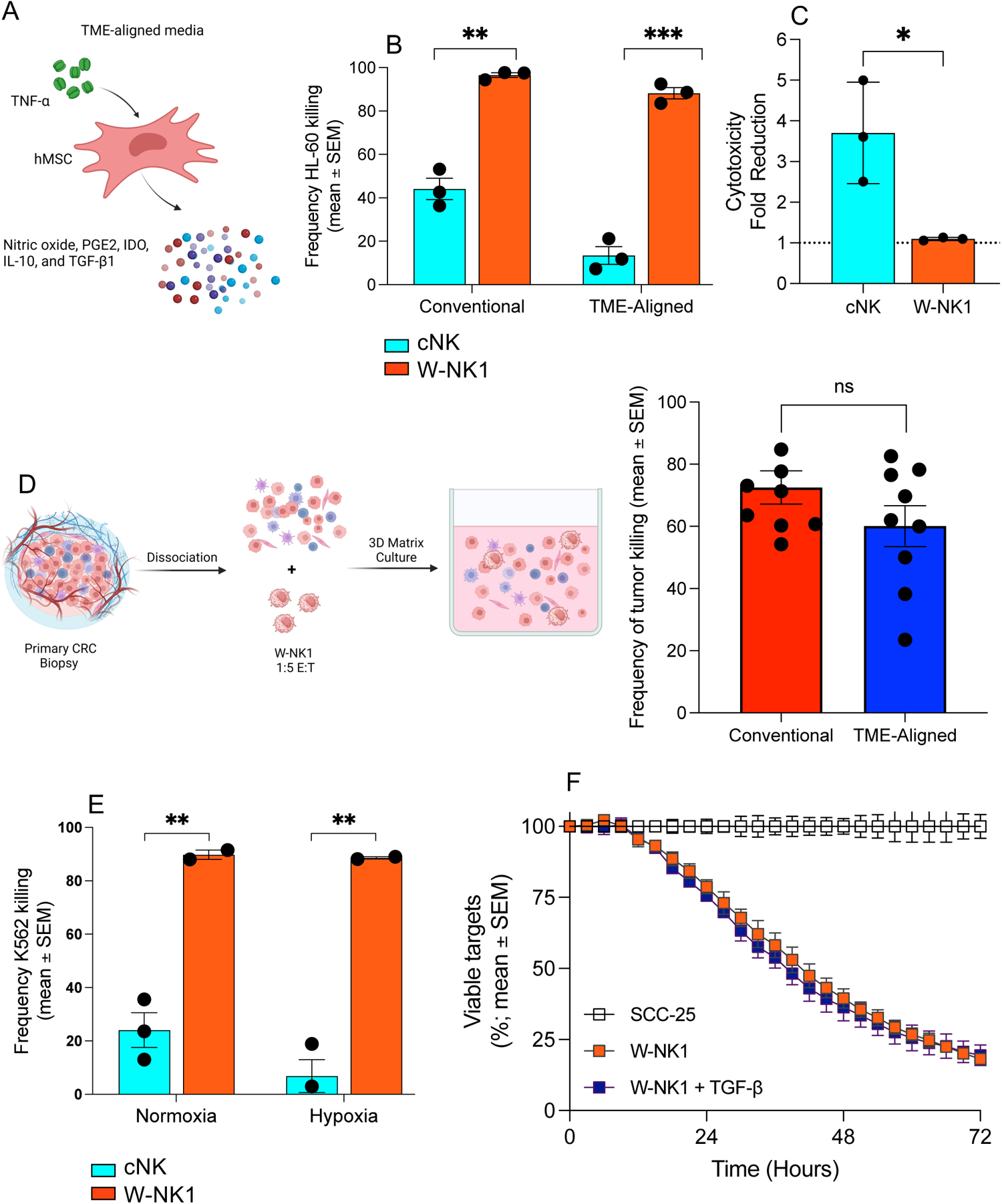
Cytotoxic activity of W-NK1 in TME models. (A) Schema depicting our approach to generating hypoglycemic (6.0 mM glucose) and acidic (pH 6.9) TME-aligned media from mesenchymal stromal cells (MSCs) stimulated with TNF-γ. (B) Effector cell cytotoxicity against HL-60 target cells in conventional or TME-aligned media expressed as frequency of target cells killed compared to no-effector control. Mean frequencies were compared by paired T test corrected for multiple comparisons, comparing conventional and TME-aligned media for each effector cell. (C) Cytotoxicity fold reduction calculated as frequency HL-60 killing in conventional media divided by frequency HL-60 killing in TME. Data are presented as mean ± SEM (n=3). Mean fold reductions were compared by a paired t-test. (D) Experimental schema. Cells from a malignant biopsy sample (colorectal carcinoma; CRC) were disassociated and embedded in BME matrix to create a 3D culture system where HL-60 target cells were incubated with W-NK1 or cNK for 48 hours with either conventional or TME-aligned media. Frequency of HL-60 target cell killing by W-NK1 after 24-hour incubation. (E) Frequency of K562 killing by W-NK1 or cNK in normoxic (5% CO_2_, atmospheric O_2_) or hypoxic (5% CO_2_, 1% O_2_) conditions normalized to live cells remaining in the ‘target-only’ control condition. Statistical difference of mean frequencies was determined by t-test, comparing cNK and W-NK1 in each culture condition. (F) W-NK1 and human head and neck tumor cell line, SCC25, were co-cultured in the absence or presence of 10 ng/mL TGF-β, a potent immunosuppressive mediator that can impair the cytotoxic ability of NK cells. The viability of the tumor cells was monitored over a 72-hour period using the IncuCyte S3 Live Cell Analysis System, with images collected every three hours. Data are expressed as % of target with mean ± SEM; n=5 replicates.

As shown in figure 4B, W-NK1 exerted significantly higher cytotoxicity against HL-60 AML targets compared to cNK in both CM (96.5% vs. 44.1%, *P = 0.006*) and TME-aligned media (88.2% vs. 13.4%, *P = 0.0006*). Importantly, W-NK1 challenged in TME-aligned media showed no change in cytotoxicity compared to CM. Conversely, TME-aligned media significantly impaired cNK-cell cytotoxicity (average = 3.7; *P = 0.02*, figure 4B). This finding was further validated using malignant patient-derived ascites in the co-cultures (figure S5). Whereas cNK-cell cytotoxicity was suppressed under the above TME conditions, W-NK1 maintained a high cytotoxic potential regardless of the culture media employed. This observation underscores an intrinsic resistance to the metabolic constraints imposed by the TME and was corroborated by native TME-aligned 3D assays with primary surgical tumor samples. In these experiments, no difference in W-NK1’s cytotoxicity was detected when comparing CM and TME-aligned media (figure 4C).

Additionally, the effects of hypoxia and transforming growth factor beta (TGF-β), a potent immunosuppressive mediator frequently secreted by cancer cells as a strategy to evade immune surveillance,^31^ on W-NK1 function were evaluated against the K562 target line in a 24-hour killing assay. Under normoxic conditions (21% O_2_), W-NK1 exhibited higher cytotoxicity against K562 cells compared to cNK (figure 4D). When exposed to a low-oxygen environment (0.1% O_2_) for 24 hours, W-NK1 maintained its ability to kill K562 cells, in stark contrast to cNK cells which exhibited reduced effector functions (figure 4D). Finally, a 72-hour killing assays conducted with and without 10 ng/ml TGF-β indicated that W-NK1’s killing of SSC-25 cells remains unaffected in the presence of exogenous TGF-β (figure 4E).

### W-NK1 exhibits increased metabolic fitness and adaptability in nutrient-deprived media

To identify metabolic features that would enable W-NK1 to exert anti-tumor cytotoxicity in nutrient-deprived TMEs, we measured the expression of cell surface nutrient transporters by flow cytometry as a proxy for W-NK1 metabolic fitness (table S4). At baseline, W-NK1 displayed a unique metabolic signature distinct from that of cNK cells and consisting of higher surface levels of glucose (GLUT1), lactate/pyruvate (MCT1), neutral amino acid (ASCT2) and inorganic phosphate (PiT1) transporters (figure 5A). After co-culturing with patient-derived malignant ascites and TME-aligned media, W-NK1 displayed increased expression of several nutrient transporters compared with conventional media, in stark contrast to cNK cells, whose protein expression profile remained largely unaltered (figure 5B). In this respect, GLUT1, which was highly expressed by W-NK1 at baseline, was unchanged in co-cultures containing ascites or TME-aligned media, whereas ASCT2, and PiT1 were upregulated when W-NK1 was maintained in either TME-aligned media or ascites. Additionally, MCT2 was also upregulated when W-NK1 was cultured in the presence of malignant ascites (figure 5B). These results support the ability of W-NK1 to source different substrates for ATP manufacturing.

**Figure 5.**
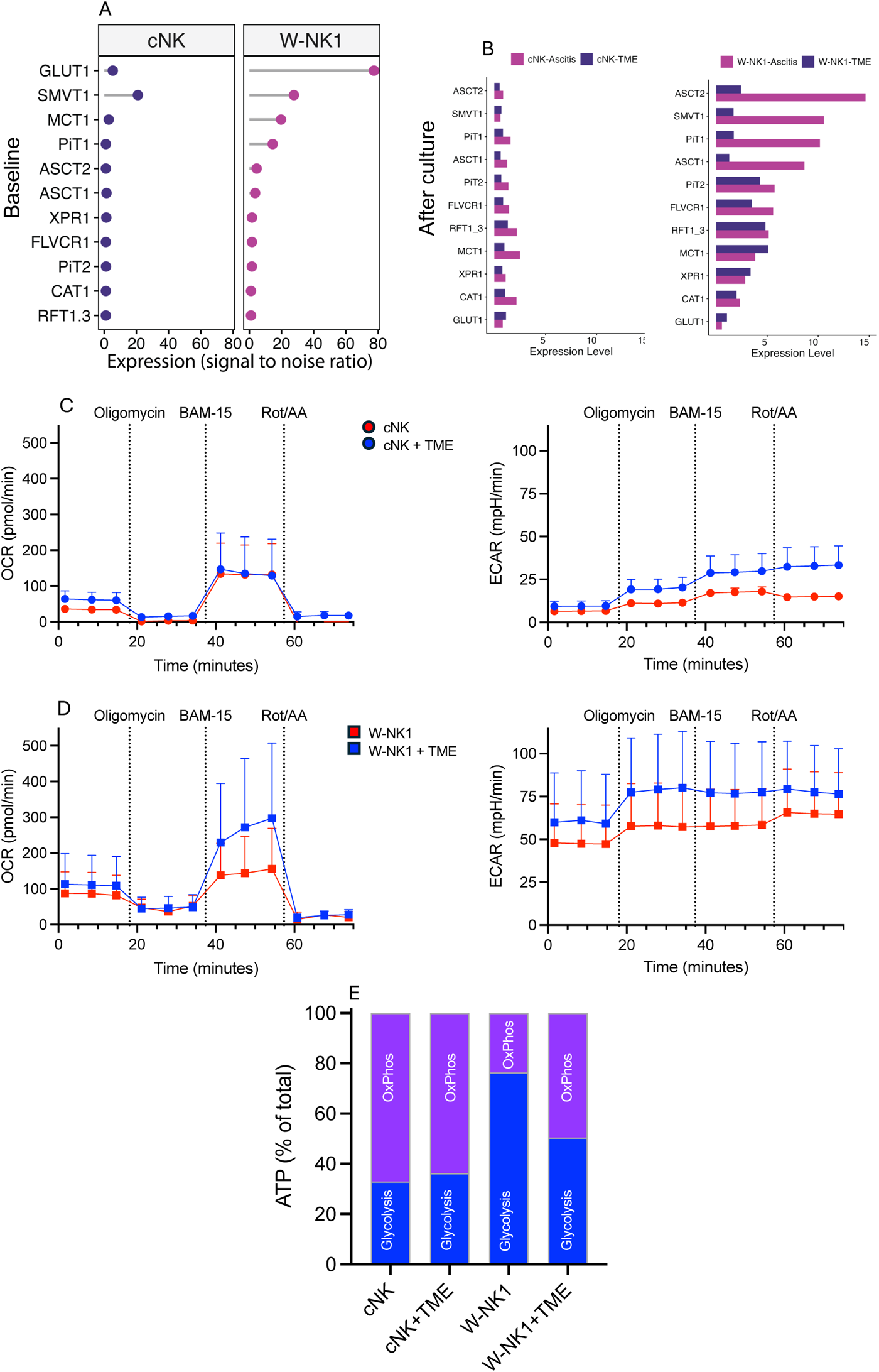
Evaluation of W-NK1’s metabolic phenotype and activity. (A) Flow cytometry evaluation of surface expression of nutrient transporters at basal level on W-NK1 compared to cNK. Expression was calculated as signal-to-noise ratio, where signal is defined as the geometric mean of the fluorescence obtained by RBD binding and noise is estimated by the fluorescence minus one (FMO) control. (B) Modulation of cell surface nutrient transporters after culture. The bar graph depicts the fold change in the expression levels of transporters in the presence of TME or patient-derived ascites compared to conventional media after 4 hours of *in vitro* culture. (C-D) W-NK1’s metabolic activity as evaluated using the Agilent Seahorse T Cell Metabolic Profiling Kit in the presence of TME-aligned culture media. The graphs show complete extracellular acidification rate (ECAR), reflecting anaerobic glycolysis, and oxygen consumption rate (OCR), an estimate of oxidate phosphorylation, from three separate experiments (three independent blood donors). (E) ATP production by W-NK1 and cNK cells through glycolysis and oxidative phosphorylation under the culture conditions detailed above.

We next conducted a Seahorse real-time analysis of W-NK1’s metabolic phenotype. This approach simultaneously measures oxygen consumption rate (OCR), an estimate of oxidative phosphorylation, and extracellular acidification rate (ECAR), which reflects anaerobic glycolysis. The OCR did not differ when cNK cells were cultured in the presence of TME-aligned media compared to CM, both at steady state and after exposure to metabolic modulators (figure 5C). In contrast, W-NK1 showed higher OCRs across time in the presence of TME-aligned media relative to CM, suggesting increased mitochondrial respiration under adverse culture conditions (figure 5D). As expected, OCR levels decreased after treatment with rotenone and antimycin-A, which inhibit the electron transport chain and are used to determine the degree of non-mitochondrial oxygen consumption. The ECAR was higher at baseline in W-NK1 cultures and increased further in TME-aligned media, indicating W-NK1’s ability to exploit multiple energy sources under stress conditions (figure 5D). In keeping with this observation, W-NK1 utilized glycolysis for 76% of its ATP production, whereas glycolysis accounted for only 32% of the ATP generated by cNK (figure 5D). Taken together, these experiment suggest that W-NK1 could maintain its metabolic fitness in the TME.

### W-NK1 shows *in vivo* trafficking and killing abilities

In order to ascertain W-NK1’s killing ability in vivo, we assessed its efficacy and biodistribution in immunodeficient (NCG) mice engrafted with human AML (THP-1 cells). In this model, W-NK1 given either as a single-dose or as three consecutive doses 7 days apart efficiently controlled AML growth compared to expanded cNK cells (figure 6A). As a single dose, W-NK1 significantly reduced tumor burden, as indicated by tumor growth inhibition (TGI) of 66.7% compared to vehicle-treated and expanded cNK-treated controls. As a multiple dose therapy, 3 weekly administrations of 1×10^7^ cells (totaling 3×10^7^ cells) enhanced W-NK1’s anti-tumor activity compared to vehicle-treated animals in terms of both tumor control (TGI = 89%) and a trend toward improved survival compared to a single dose of 1×10^7^ cells (figure 6A). Given our observation that W-NK1 has increased expression of tissue homing chemokine receptors, such as CXCR4, CXCR3, CCR7 and CCCR5, we evaluated its ability to migrate to the BM as well as to extramedullary sites *in vivo*. Indeed, in non-tumor bearing mice, W-NK1 efficiently homed to the BM as well as the lung and liver within 24h of i.v. injection (figure 6B). While the pattern of W-NK1 trafficking was similar to that previously reported for other NK cells, homing to the BM was ∼3 fold higher (1-5% *vs.* 16%).^32^

**Figure 6.**
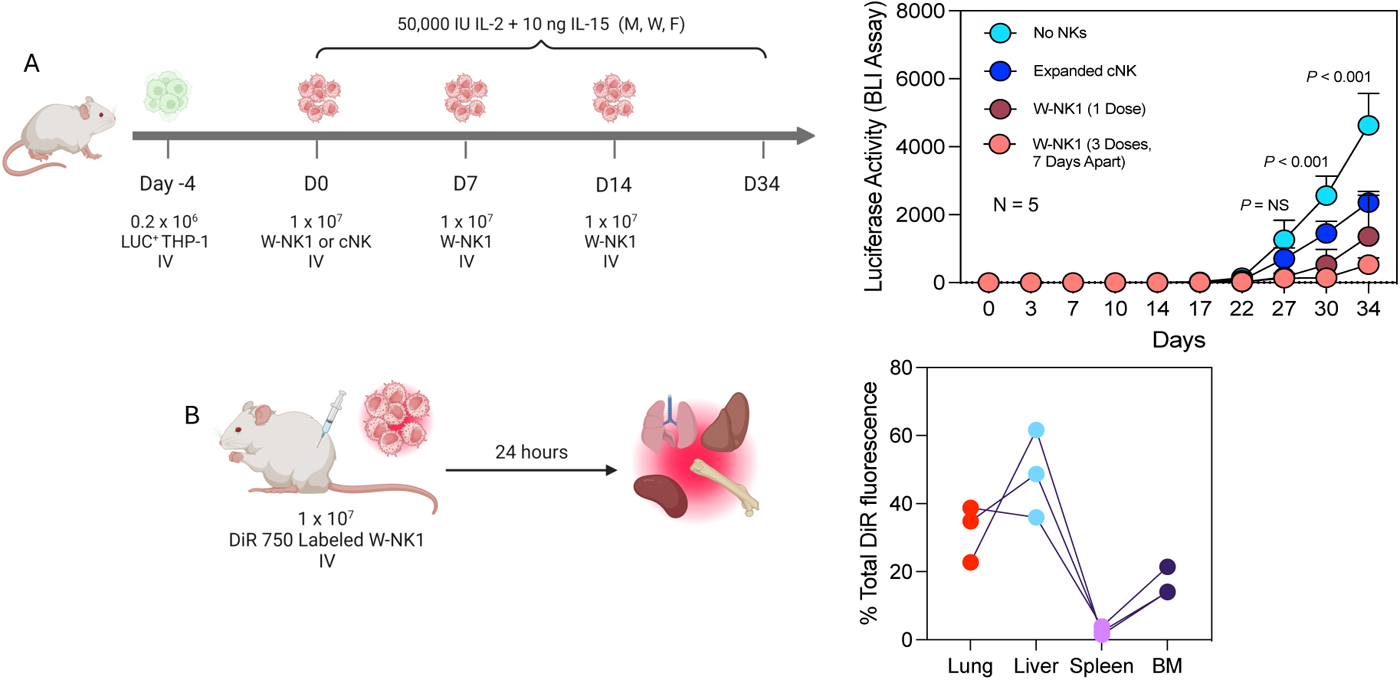
W-NK1 *in vivo* trafficking and cytotoxicity. (A) Experimental schema and THP-1 tumor growth measured by luciferase activity. Briefly, luciferase-expressing THP-1 cells were injected i.v. into NSG mice at 0.2×10^6^ cells/mouse four days before NK infusion. Five mice per group were injected with a single i.v. bolus of expanded cNK (1×10^7^), W-NK1 (1×10^7^), or three doses of W-NK1 (1×10^7^) at weekly intervals. A control group received vehicle only. THP-1 growth was monitored by whole-body bioluminescent imaging (BLI) every 3-4 days to day 34 post NK treatment. (B) Biodistribution of W-NK1 (n = 3) labeled with IVISense DiR 750 Fluorescent Cell Labeling Dye, detected by fluorescent signal of excised organs as a percentage of total signal detected in these organs of each mouse.

### W-NK1 expands and differentiates in a patient with AML

We finally evaluated the *in vivo* phenotypic changes of W-NK1 in one patient with AML enrolled in study NCT05470140, taking advantage of HLA antigen disparity between the donor (drug product [DP], W-NK1) and recipient (patient) cells. This ongoing trial recruits patients with relapsed or refractory AML who have exhausted all other approved therapies receive W-NK1 on days 1, 8 and 15 of a 28-day cycle, at three dose levels ranging from 300 to 1,800×10^6^ cells per infusion during dose escalation (figure 7A). The patient reported herein, who met the HLA-disparity criteria, was in their 70s and had a history of relapsed AML, adverse risk cytogenetics^33^ and a baseline disease burden of 50% BM blasts prior to entering the study. After failing multiple prior lines of therapy (cytarabine and daunorubicin (7+3), fludarabine, cytarabine, idarubicin, and granulocyte colony-stimulating factor (G-CSF) (FLAG-Ida), high dose cytarabine (HiDAC), venetoclax and azacytidine, venetoclax, low-dose cytarabine and cladribine (Ven/LDAC/Cladribine), and FLAG-Ida in combination with venetoclax), the patient received W-NK1 at dose level (DL) 2 (900×10^6^ cells per infusion). Pharmacokinetic and pharmacodynamics analysis (figure 7B) demonstrated approximately a 4-log expansion of W-NK1 *in vivo*, with persistence through day 14, with concurrent effective elimination of circulating AML blasts. Interestingly, BM biopsy of D28 shows increased staining of CD56 compared to the pre-treatment biopsy, suggesting W-NK1 trafficking and persisting in the BM following expansion in the peripheral blood (figure 7D). Analysis of W-NK1 phenotype in the peripheral blood (supplemental figure 6) showed continued evolution, adaptability, and maturation. W-NK1 was detected in patient samples by flow cytometry analysis and showed peak expansion at day 10 of the treatment cycle, consistent with pharmacokinetic analysis (figure 7C). The phenotype of post-infusion W-NK1 was then compared to that of the drug product before administration (DP) and donor-matched cNK. The patient’s endogenous lymphocytes were identified by HLA Bw4 staining, which was absent on W-NK1 (figure S7).

**Figure 7.**
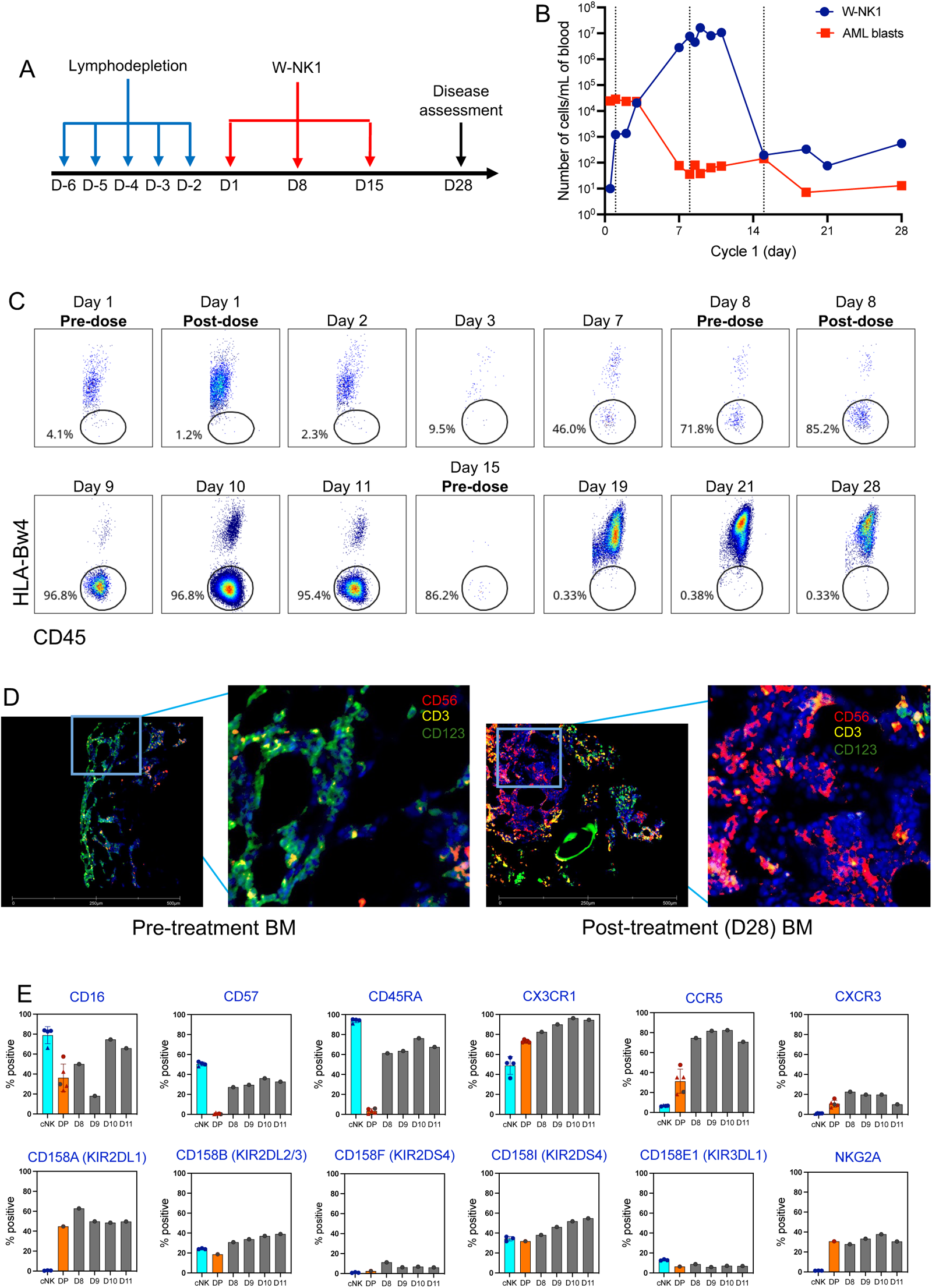
Detection of W-NK1 in a patient with acute myeloid leukemia. (A) Clinical study schema. (B) Pharmacokinetic and pharmacodynamics of W-NK1 measured by SNP variable allele frequency (VAF) and normalized to cell count. Vertical dashed lines indicate timing of NK-cell dosing. (C) Frequency of HLA-Bw4-negative W-NK1 in the patient’s peripheral blood (PB) at the indicated timepoints (pre and post W-NK1 infusion). (D) Bone marrow FFPE biopsy pre and post (Day 28) dosing with W-NK1 (green: CD123 as a marker for AML blasts, red: CD56 as a marker for NK cells, yellow: CD3 as a marker for T cells). (E) Percent positive cells for the indicated markers (donor cNK, DP and patient’s PBMCs) at different timepoints (D8-D11).

Phenotypic analyses of the DP *in vivo* showed that, compared to the pre-infusion DP, post-infusion W-NK1 acquired a more mature phenotype with increased expression of CD45RA and CD57. We also detected phenotypic changes suggestive of improved homing with an increase in the expression of CXCR3, CX3CR1 and CCR5, which, beyond homing, have been shown to be essential for improved anti-tumor activity of NK cells.^17,33^ Additionally, we observed expression of key receptors mediating cytotoxic function, *i.e.*, CD16, NKp44, NKp80, NKp46, 2B4 and KIRs (figure 7E), consistent with NK-cell education and licensing.^34,35^ These observations illustrate that W-NK1 with a matures, expands and persists in patients with AML.

## DISCUSSION

Ideal features for successful ACT that have emerged from chimeric antigen receptor (CAR) T-cell clinical trials include 1) tumor-targeting selectivity and specificity to minimize toxicity and prevent antigen escape or acquired resistance; 2) efficient trafficking and infiltration into tumors; 3) robust anti-tumor activity and 4) resilience to adverse and immunosuppressive TMEs.^12,36,37^ There is urgent need to develop cellular products that potentially overcome the limitations of ACT. In this respect, genetic engineering of innate-like T lymphocytes and NK cells translates into functional reprogramming, leading to enhanced anti-tumor activity and prolonged *in vivo* persistence.^38^ In particular, NK cells are ideally positioned to improve immunotherapy outcomes, in light of their safety profile,^39^ ability to eliminate tumor cells with down-regulated MHC-I expression, orchestrate adaptive immune responses and expand in response to cytokine stimuli.^4^

Our study offers a comprehensive characterization of the phenotypic and functional attributes of W-NK1, a cytokine-primed and expanded ML NK cell therapy product. Our multi-modal omics approach revealed significant upregulation of genes, proteins, and metabolites involved in glycolysis, amino acid metabolism, and fatty acid oxidation in W-NK1. These pathways were consistently and reproducibly identified across transcriptomic, proteomic, and metabolomic datasets, underscoring the robustness of our findings and emphasizing W-NK1’s therapeutic potential in challenging TMEs. When compared to the pre-manufacturing cNK-cell population, W-NK1 demonstrated enhanced cytotoxicity against AML and other malignant cell lines, thereby outperforming cNK cells while showing no indication of off-target effects in pre-clinical models, and in contrast to other immunotherapy modalities including CART cells and bispecific antibodies.^40^ Unlike cNK cells, W-NK1 sustained high cytotoxic potential and intrinsic resistance to metabolic constraints imposed by the TME, including nutrient deprivation, hypoxia, and high concentrations of immunosuppressive mediators.

W-NK1’s enhanced resilience could be related to its increased metabolic flexibility, which is suggested by the dynamic expression of nutrient transporters and the ability to switch between glycolysis and oxidative phosphorylation depending on nutrient availability and oxygenation levels, as also suggested by previous studies.^41–44^ Similarly, cytokine-induced upregulation of fatty acid oxidation has been reported to promote NK-cell cytotoxicity.^45^ The characteristics of W-NK1’s metabolic behavior reported in our study are in keeping with the principles of Warburg metabolism, which underpins the upregulation of glycolysis even in the presence of oxygen. Cells benefit from Warburg metabolism by maintaining a reservoir of glycolytic intermediates, which supports rapid cell proliferation and various biosynthetic pathways.^20^ In this respect, W-NK1’s reliance on Warburg metabolism may provide a diverse pool of energy substrates essential for its cytotoxic function, particularly for the energy-intensive process of serial target lysis.

Ineffective homing of ACT to tumor sites has been linked to off-target toxicity and reduced clinical activity.^46^ Accordingly, efforts to enhance CART-cell homing to the BM have translated into better outcome in AML.^47^ Our pre-clinical models highlighted W-NK1’s ability to migrate to the BM, leading to effective tumor control. Importantly, multiple doses of W-NK1 enhanced its anti-tumor activity and improved mice survival. Phenotypic analysis in one patient with relapsed AML who received W-NK1 therapy revealed increased expression of homing receptors and key cytotoxicity molecules on the ‘drug product’ (post-manufacturing W-NK1), with further upregulation post-infusion. By analyzing BM sections and serial blood samples, our study also provides evidence for W-NK1’s *in vivo* persistence and phenotypic evolution, including a more mature phenotype and increased homing and cytotoxicity receptor expression post-infusion. Importantly, W-NK1’s peak expansion on day 10 post-infusion was concurrent with a reduction of circulating AML blasts. Collectively, these findings underscore the concept of W-NK1 as a “living drug” which could continuously evolve its molecular and functional features along a continuum of phenotypic and functional states, spanning cNK cells as the starting material, pre-infusion cytokine-primed and expanded W-NK1, and post-infusion W-NK1. W-NK1 is currently being evaluated in both hematological (NCT05470140) and solid tumor malignancies (NCT05674526). Future research should also focus on exploring its potential in combination with other therapeutic modalities.

## MATERIALS AND METHODS

### Generation of W-NK1

Previously frozen conventional NK cells (cNK) derived from allogeneic healthy donor whole blood were reprogrammed and expanded through a GMP-grade cytokine complex feeder cell-free process and cryopreserved using a proprietary method (MONETA™) to get an off-the-shelf NK-cell product (W-NK1).

### scRNA-sequencing

We profiled the transcriptome of four batches of W-NK1 (derived from different donors) at single-cell resolution using the Chromium Next GEM Single Cell 3’ Reagent Kit v3.1 (10X Genomics). Raw FASTQ files were processed using Cell Ranger (v7.1.0) to map reads against human genome 38 as a reference, filter out unexpressed genes, and count barcodes and unique molecular identifiers (UMIs). We retrieved 64,973 cells over 8 samples, with 31,878 detected genes (87.1%). After identifying adaptive, sample-specific thresholds to eliminate low-quality cells (*scater* package in R), filtering out blacklisted genes (including *MALAT1*, hemoglobin, mitochondrial, ribosomal and immunoglobulin) as well as genes based on sparsity (i.e., genes detected at low levels in a small number of cells), and excluding doublets (DoubletFinder package in R), we retained 54,453 cells for downstream analyses with the Seurat R package (version 5.0). To mitigate unwanted technical variation and address potential confounders, we normalized the dataset using SCTransform, setting the method parameter to glmGamPoi, invoking the v2 regularization, and passing the cell cycle difference (defined as the difference between S phase and G2M phase module scores calculated using the *CellCycleScoring* function) to the *vars.to.regress* argument. We conducted data integration by selecting the top 3,000 variable features and identifying anchors between the datasets (cNK and W-NK1) with the *FindIntegrationAnchors* function (k.anchor = 20, reduction = ‘rpca’, dims = 1:30), followed by data scaling and PCA.

Initially, from the first-run clustering, contamination clusters expressing signature genes associated with monocytes, T cells and B cells were identified and removed using SingleR (MonacoImmuneData). The cleaned dataset comprised 49,203 high-quality NK cells (23,829 cNK and 25,374 W-NK1). Subsequently, the data was re-normalized using the SCTransform package in R, followed by the PCA-UMAP clustering pipeline in Seurat with a resolution of 0.2. For the second-run clustering, the top 30 PCs based on 2,000 highly variable genes were selected (resolution = 0.2). Differentially expressed genes identified in Seurat (*FindAllMarkers* function) using the limma implementation of the Wilcoxon rank sum test^48^ (adjusted *P* < 0.01; logFC threshold = 0.25; minimum percentage of cells expressing the gene greater than 25% in at least one of the compared clusters) were chosen for manual cluster annotation (table S2), resulting in 8 cellular states (c0-c7). Gene signature scores were calculated using the *AddModuleScore* function, which estimates the average expression of each gene program at the single-cell level, after subtracting the aggregated expression of a control feature set. Single-sample GSEA (ssGSEA) on sc-RNA-seq data was conducted using the escape package in R, employing the Hallmark gene set (*UCell* method).^49^ Trajectory graphs of gene expression changes after cytokine priming were built using the learn_graph function in Monocle3, which is a diffusion pseudotime algorithm that learns the sequence of gene expression changes and identifies developmental branch points.^50^ Pseudotime was inferred with the order_cells function.

### Multi-omics, in vitro and in vivo functional assays, and immunofluorescence staining of primary BM samples

Details are provided in Supplementary Materials and Methods.

### Statistical analyses

Descriptive statistics included calculation of median, inter-quartile ranges and proportions to summarize study outcomes. Comparisons were performed with the Mann-Whitney *U* test for paired or unpaired data (two-sided), as appropriate, or with the ANOVA with correction for multiple hypothesis testing (Benjamini-Hochberg procedure). A *P* value less than 0.05 was considered significant. IBM SPSS Statistics (version 29), R (version 4.4.0) and GraphPad Prism (version 10.2.3) were used for statistical analyses and/or to generate figure plots.

## Contributors

JV, JM, JKDM, and SR conceived of, designed, and supervised the study; LA, NM, JV, TL, DJB, CC, AH, KM, JD, ES, BC, VP, NB, CP, BT, LW, LL, YHR, AMM, PCP, DP, ND, SKL, NE, JM, JKDM, and SR performed the experiments and/or analyzed the data; AY and CD performed the Seahorse experiments; LA, NM, and SR wrote the original draft; JV, DB, MMBE, TAF, JKDM, and SR reviewed and edited the paper. SR is responsible for the overall content as the guarantor.

## Acknowledgements

The authors are grateful to Professor Karl-Johan Malmberg and Mr. Herman Krogstad Netskar, Oslo University Hospital, Norway, for granting early access to the processed R object and related annotation for the NK-cell reference map, and for insightful discussions.^27^

## Ethics approval

This study has received approval from the Institutional Review Board of WCG (Western Copernicus Group), Princeton, NJ (Study Number: 1352242; IRB tracking number: 20226001). Participants gave informed consent to sample collection, which was performed as part of their routine clinical care.

## Data availability statement

The mass spectrometry proteomics data have been deposited to the ProteomeXchange Consortium *via* the PRIDE partner repository with the dataset identifier PXD052855. The newly generated bulk RNA-seq and sc-RNA-seq datasets have been deposited to Zenodo as a row count matrix and a processed Seurat object, respectively (10.5281/zenodo.11441148). Source data needed to evaluate the conclusions in the paper are provided in the paper and/or the Supplementary Materials.

---

**Figure S1 (Related to Fig. 3).**
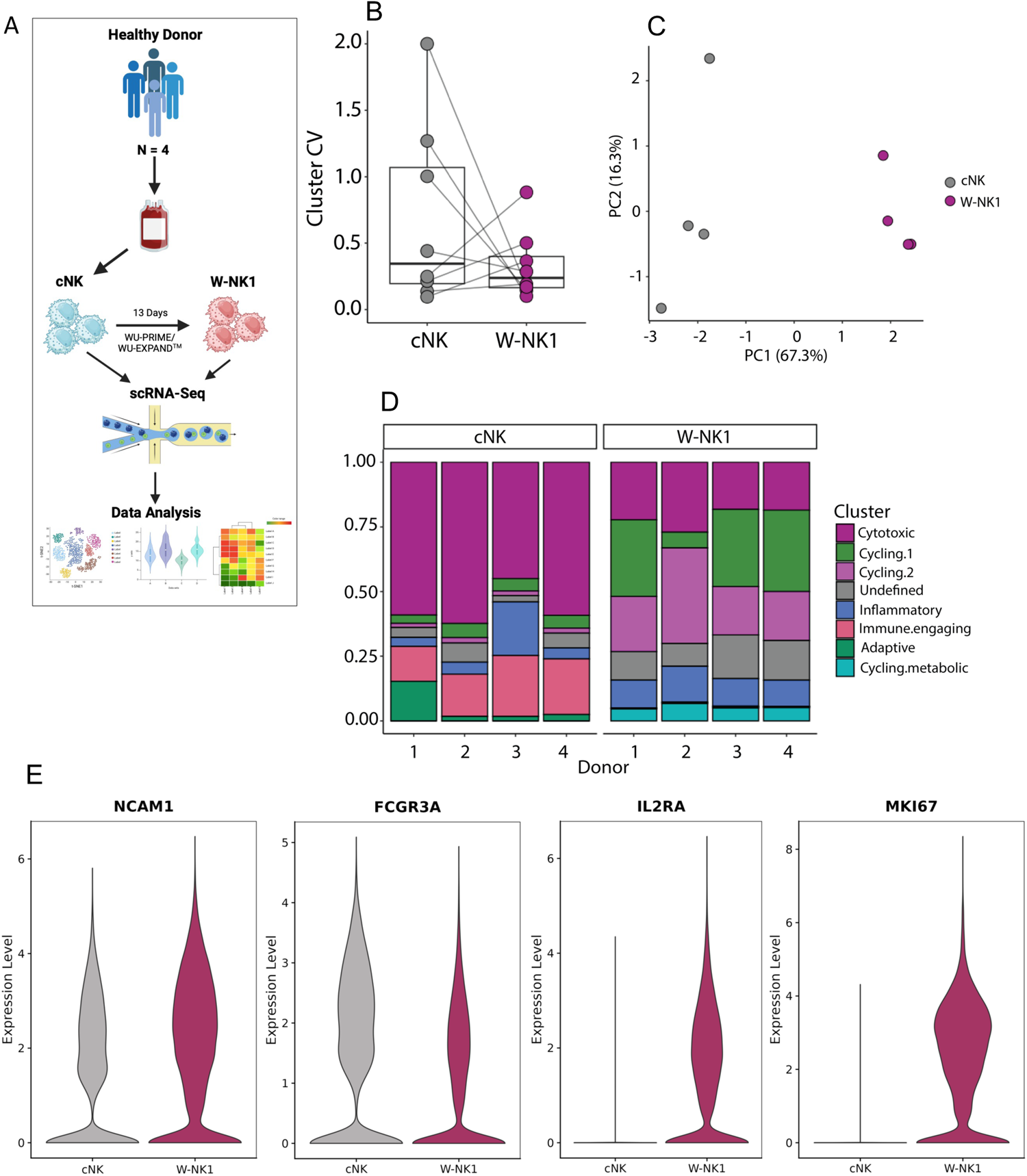
Single-cell transcriptomic analysis of W-NK1. (A) Schematic representation of the experimental design. (B) Variance analysis of sc-RNA-seq clusters. The box plot shows the distribution of cluster coefficients of variation (CV) for W-NK1 and cNK cells. Each point represents the CV of an individual cluster, with lines connecting the corresponding clusters between the two groups. The *P* value represents the result of a paired t-test. (C) PCA plot showing the distribution of cluster frequencies in W-NK1 (magenta) and cNK cells (grey). Each point represents an individual sample, positioned according to its principal component scores. PCA was performed on frequency data of cell clusters, scaled and centered prior to analysis. (D) Stack bar plot summarizing compositional differences between cNK and W-NK1 across four paired samples. (E) Violin plots showing NCAM1, FCGR3A, IL2RA and MKI67 expression in W-NK1 and cNK cells.

**Figure S2 (Related to Fig. 3).**
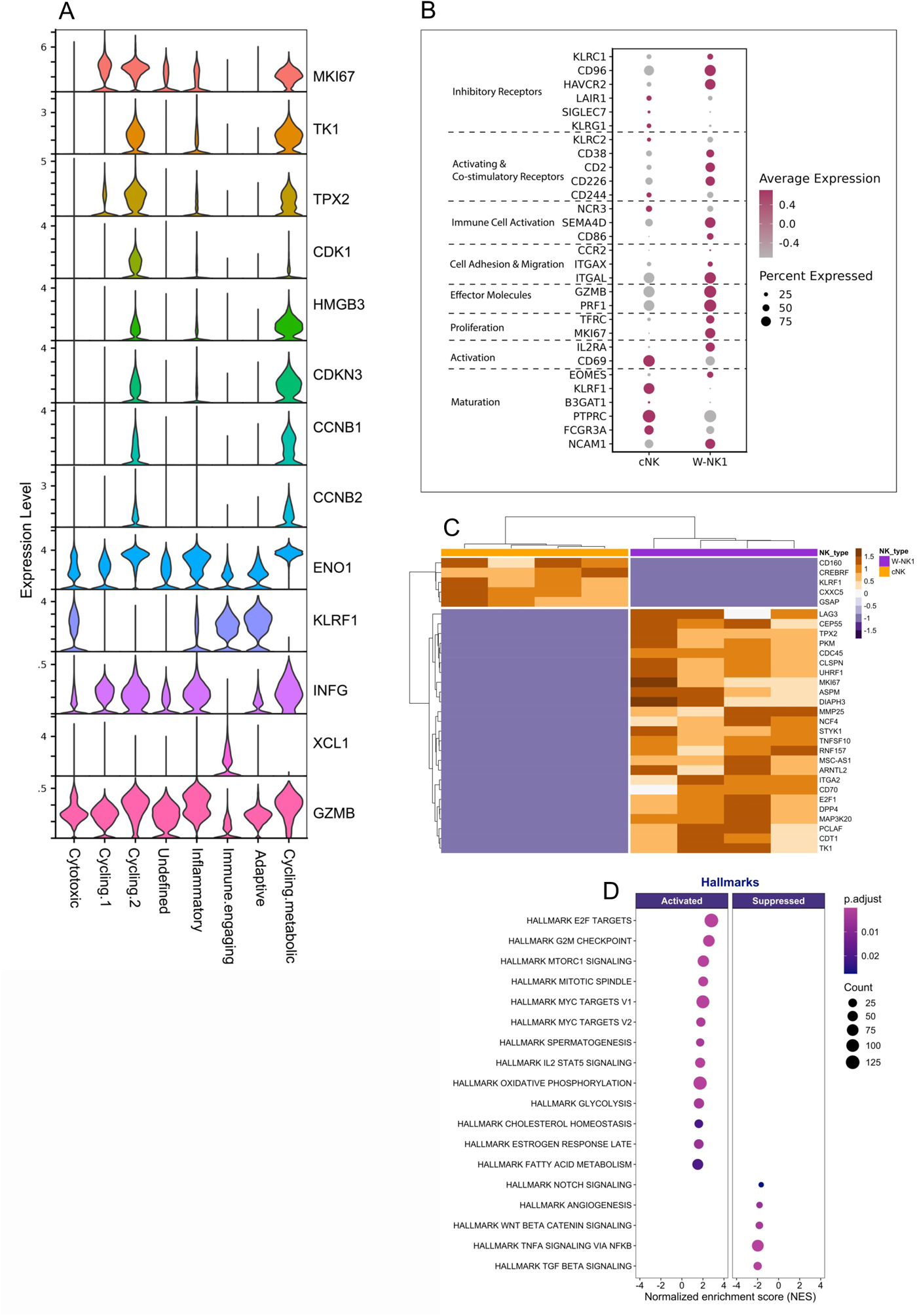
Single-cell transcriptomic analysis of W-NK1. (A) Violin plots depicting the expression of key selected markers discussed in the main text. (B) Dot plot showing the expression of key NK genes grouped by their known functional role in NK biology. (C) Heatmap summarizing the expression of the top 30 differentially expressed genes (DEGs) in cluster 0 cells between W-NK1 and cNK. Genes were identified using pseudobulk analyses. (D) Dot plot showing the top enriched Hallmark pathways using DEGs from pseudobulk analyses of c0 cells as input (clusterProfiler package in R).

**Figure S3 (Related to Fig. 3).**
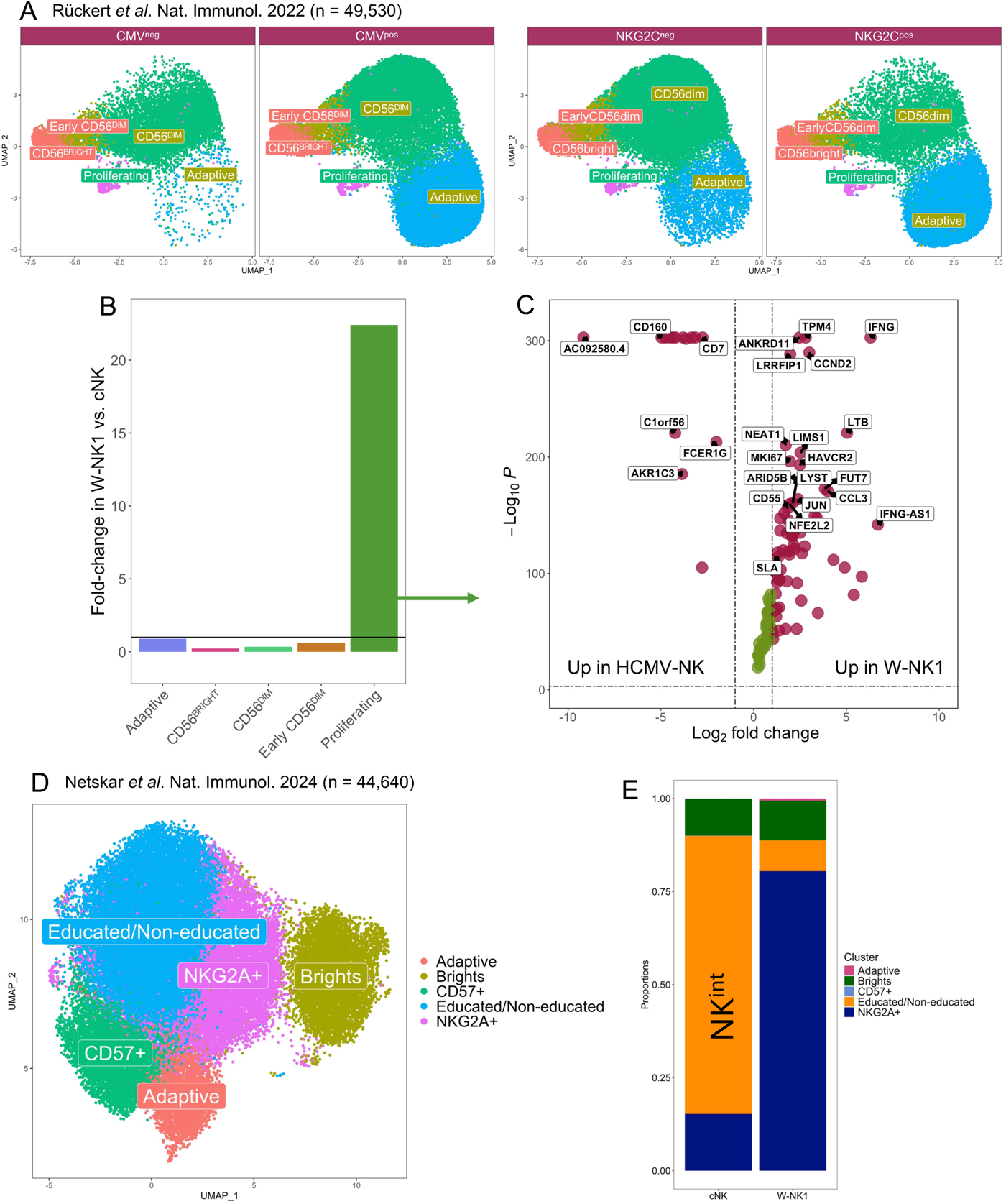
Comparison of W-NK1 to external datasets using transfer learning. (A) UMAP representation of healthy donor (HD)-derived NK cells from Rückert *et al*. (GSE197037). Cells are visualized based on CMV status and NKG2C expression. (B) Bar graph comparing cluster abundance in NK cells from HD (reference map) and W-NK1, which was annotated using label transfer with ProjecTILs. (C) Volcano plot of differentially expressed genes in proliferating NK cells from the reference map and W-NK1. (D) UMAP representation of HD-derived NK cells from Netskar *et al*. (https://zenodo.org/doi/10.5281/zenodo.8434223). (E) Bar graph showing compositional differences between W-NK1 and cNK cells (linear modeling with speckle in R) annotated using Seurat’s standard pipeline for mapping and annotating query datasets. Uneducated/educated NK cells were labeled as ‘NK^int^’ in the original manuscript (Netskar *et al*. Nat. Immunol. 2024).

**Figure S4 (Related to Fig. 5).**
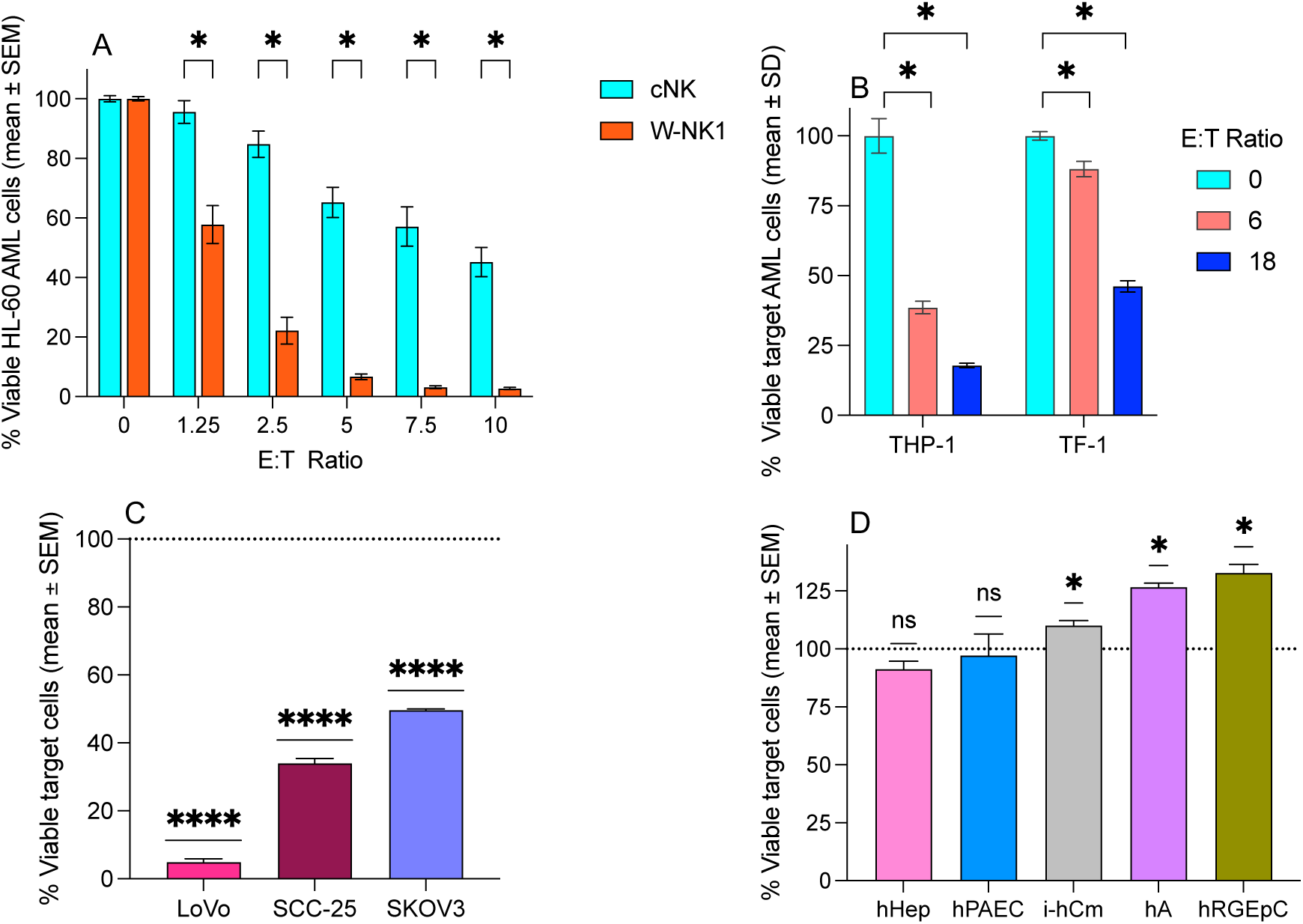
Cytotoxicity assays against malignant and non-malignant human cell lines. (A) Percentage of viable HL-60 AML cells after 48-hour incubation with paired cNK or W-NK1 at the indicated E:T ratios, normalized to the control, no NK-cell condition (E:T=0). The difference between cNK and W-NK1 for each E:T ratio was determined using a paired t-test with correction for multiple comparisons. *Adjusted *P* < 0.001. (B) Percentage of viable target AML cells after 72-hour incubation with W-NK1 at the indicated E:T ratio, normalized to the control, no NK-cell condition (E:T = 0). The difference between cNK and W-NK1 for each E:T ratio was determined using a paired t-test with correction for multiple comparisons. *Adjusted *P* < 0.001. (C) Percentage of viable target cells after 72-hour incubation with W-NK1 at 3:1 E:T ratio. A one-sample t-test was used to compare each mean to 100% viability of the target from the control culture condition without W-NK1 (E:T=0). (D) Percentage of viable target cells after 72-hour incubation with W-NK1 at 3:1 E:T ratio. A one-sample Wilcoxon test was used to compare each mean to 100% viability. *Adjusted *P* < 0.001. hPAEC = primary human pulmonary artery endothelial cells; hRGEpC = human renal glomerular epithelial cells; hA = human astrocytes; i-hCm = human iPSC-derived cardiomyocytes; hHep = human hepatocytes.

**Figure S5 (Related to Fig. 5).**
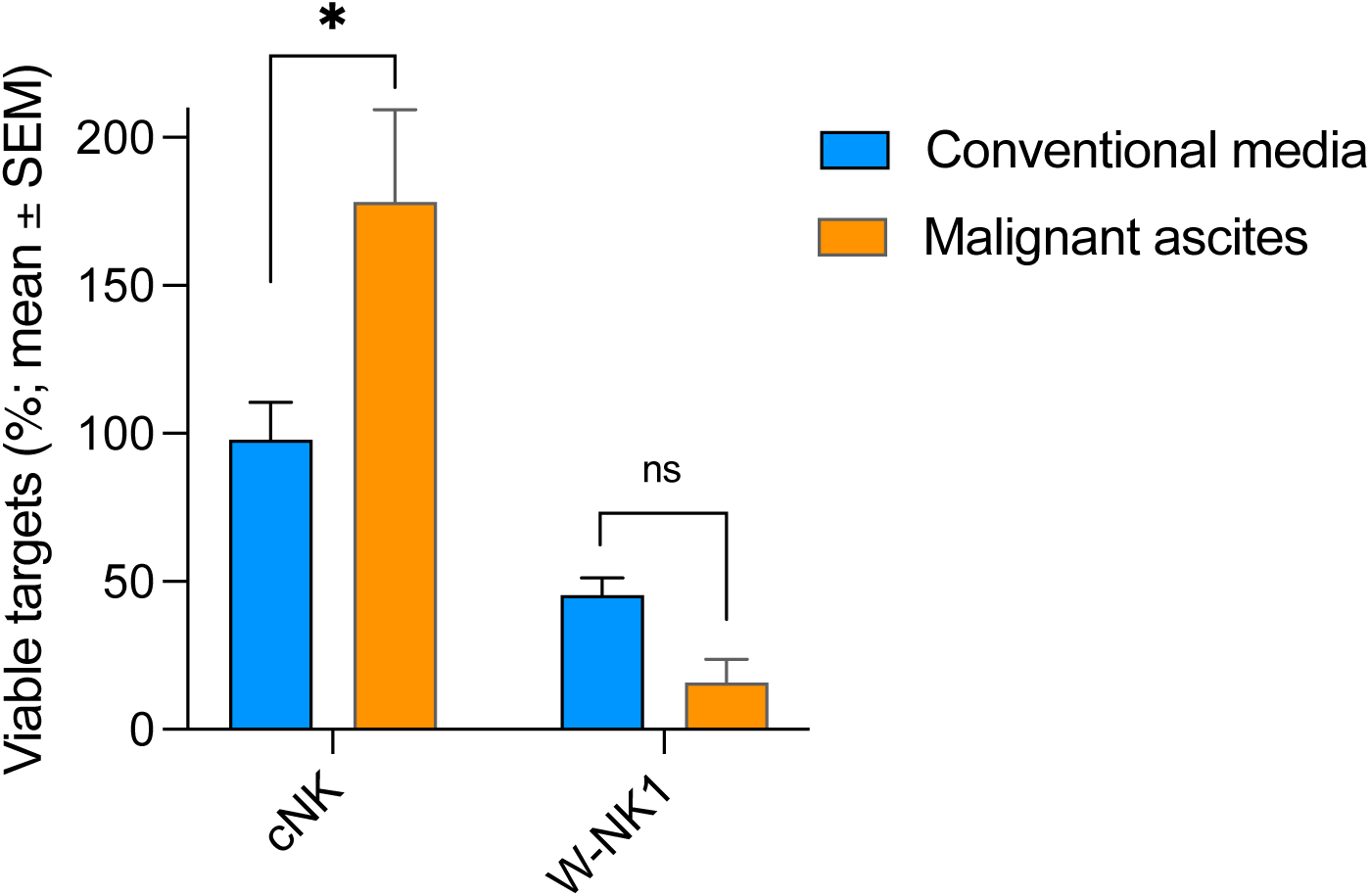
W-NK1 retain cytotoxic function in malignant ascites derived from patients with ovarian cancer. Donor-matched cNK and W-NK cells were used as effectors against HL-60 target cells (10,000 cells/well) expressing Nuclight Red at a 3:1 effector to target (E:T) ratio. Cells were incubated in either control media (DMEM with 20% FBS) or malignant ascites, both containing 100 IU/ml of IL-2. IncuCyte imaging was used to assess target cell killing. Results are graphed as percentage of viable target cells divided by viable target cells in the target only condition at t = 70 h. Results from 2 separate malignant ascites donors are pooled.

