## Supplemental Materials for "An integrated multi-omics investigation of W-NK1, a cytokine-primed non-engineered natural killer cell therapy product"

**Arthur L, *et al*.**

### SUPPLEMENTAL MATERIALS AND METHODS

#### Cell culture

For functional assays, we employed the following culture media: 1) conventional cell growth media (CM; 20% IMDM [IMDM, FBS 20%, and penicillin-streptomycin antibiotics 1x]; RPMI1640 GlutaMAX™ HEPES + 10% heat inactivated human AB serum); 2) TME-aligned media (hypoglycemic [6.0 mM glucose]; 3) acidic [pH 6.9] conditioned media obtained from stimulated human mesenchymal stromal cells (hMSCs), which are known to secrete soluble molecules, such as nitric oxide, PGE2, IDO, IL-10, and TGF-β1, with immunosuppressive functions[1] and 4) malignant ascites derived from patients with ovarian cancer (Washington University St. Louis, MO). For 3D culture assays, cells were embedded in Cultrex® Basement Membrane Extract (BME), a soluble form of basement membrane (BM) purified from Engelbreth-Holm-Swarm (EHS) tumor mimicking an in vivo TME, including low glucose and low pH. BME contains laminin, collagen IV, entactin, and heparan sulfate proteoglycan and provides a natural extracellular matrix hydrogel that polymerizes at 37ºC to form a reconstituted BM. Once solidified (approximately 3h at 37ºC), the 3D gels were covered with cell growth media (either CM or TME-aligned).

#### Bulk RNA sequencing

Total RNA was extracted from eight NK products generated from matched peripheral blood samples and used for RNA-seq analysis at the Genome Technology Access Center (Washington University School of Medicine, St. Louis, MO, USA). Briefly, samples were prepared, indexed, pooled, and sequenced on an Illumina NovaSeq 6000. Basecalling and demultiplexing were performed with Illumina's bcl2fastq2 software. RNA-seq reads were then aligned and quantitated to Ensembl release 101 with an Illumina DRAGEN Bio-IT on-premises server running version 3.9.3-8 software. For secondary analysis and visualization, gene counts were imported into R/Bioconductor packages, as detailed in the key resources table.

### Immunofluorescence staining of primary BM samples

Formalin-fixed paraffin-embedded (FFPE) BM sections obtained at baseline and on day 28 after W-NK1 infusion were prepared by deparaffinization and rehydration involving three washes in Xylene (Sigma Aldrich, Cat. No. 534056), followed by two washes in 100% ethanol (Merck, Cat. No. 1009832511), and then two additional washes in 95% ethanol. Subsequently, the tissue slides were rehydrated in double-distilled water before further processing. Antigen retrieval was performed using 1X citrate buffer (Sigma Aldrich, Cat. No. C999) within a preheated pressure cooker for a duration of 15 minutes. Following antigen retrieval, the slides were thoroughly rinsed with 1X tris-buffered saline containing 0.1% Tween-20 (TBS-T) for 5 minutes. Fc receptor blocking was accomplished by incubating the slides in 200 µL of Buffer-W at room temperature in a humidity chamber for 1 hour. After this step, the slides were incubated with unconjugated mouse monoclonal anti-CD123 antibody (NCL-L-CD123, Leica Biosystem) for one hour at room temperature, were washed three times with 1X TBST, and further incubated with Alexa Fluor™ 532-conjugated anti-mouse secondary antibody (A-11002, Thermo-Fisher) for 45 minutes at room temperature. Excess antibodies were removed by washing in 1X TBST for three times at room temperature. Sections were then incubated with AF592-conjugated anti-CD3 (NBP2-54392AF594, Novus) and AF647-conjugated anti-CD56 (NB100-2718AF647, Novus) antibodies for one hour at room temperature. Nuclei were stained with SYTO 13 (Invitrogen, Cat No. S7575). The slides were subsequently washed with 1X TBST and subjected to imaging using the NanoString GeoMx Digital Spatial Profiling platform.

### Flow cytometry

Paired cNK and W-NK1 (n = 28) were analyzed by flow cytometry (IMU Biosciences, London, UK). Samples were thawed in 37°C water bath and washed twice in thaw media (10% fetal bovine serum in RPMI) and PBS, sequentially. Cells were resuspended in PBS and cell count was estimated using SONY ID7000 spectral flow cytometer (SONY Biotechnology) prior to viability staining using Live/Dead® Fixable Blue Dead Cell Stain (Life Technologies) for 15 minutes at room temperature. Cells were then washed and resuspended in FACS buffer (2 mM EDTA and 0.5% BSA in PBS) for staining. Samples were split and stained using two antibody master mixes (NK1 and NK2; table S1). For NK1 panel, 50 µL of $0.25×10^6$ of NK cells or $1×10^6$ PBMCs were stained first with the chemokine receptor master mix (made with Brilliant stain buffer plus (BD) and TruStain Fc receptor blocker (BioLegend) in 37°C water bath for 15 minutes prior to 30-minute surface staining at room temperature with remaining surface marker master mix (including Brilliant stain buffer plus (BD)). Cells were then fixed using BD lysing buffer (BD) and washed once with FACS buffer prior to acquisition on SONY ID7000. For NK2 panel, 100 µL of $0.5×10^6$ of NK cells or $2×10^6$ PBMCs were incubated with surface marker master mix (made with Brilliant stain buffer plus (BD) and TruStain FcX Receptor Blocking Solution (BioLegend) for 30 minutes at room temperature and washed with FACS buffer before fixing and permeabilizing cells for 45 minutes using fixation/permeabilization reagent (Invitrogen™ eBioscience™ Foxp3 / Transcription Factor Staining Buffer Set) prepared according to manufacturer's instructions. Permeabilized samples were washed twice with perm buffer, incubated for 45 minutes with the intracellular marker master mix and washed twice again with perm buffer before acquisition on SONY ID7000. The same cytometer, with the same configuration, was used for the measurements of all the samples. AlignCheck particles (SONY) and 8 peak beads (SONY) were run daily to perform instrument QC and to standardize the cytometer using median fluorescence intensity (MFI) target values, respectively. Flow cytometry data (FCS files) were processed and analyzed using the Dotmatics OMIQ online platform. The intensity of marker expression was normalized through arcsinh transformation. Cellular debris and doublets were excluded based on forward scatter (FSC) and side scatter (SSC) characteristics. Additionally, dead cells were removed from the dataset using Live/Dead Fixable Blue staining. For the identification of immune cell subpopulations, we employed unsupervised clustering on the live cell population using the FlowSOM algorithm, which was executed with its default settings. Subsequent dimensionality reduction to visualize the data was performed using the Uniform Manifold Approximation and Projection (UMAP) algorithm. The markers utilized in both the FlowSOM and UMAP analyses are detailed in table S1. The expression of lineage markers (CD45, CD56, CD14/CD19, CD3, CD8) facilitated the identification of myeloid and T-cell clusters, which were excluded from further analyses. Median expression and frequency of NK cells positive for each marker was compared in cNK and W-NK1 samples by unpaired t-test.[2]

#### Flow cytometry-based metabolic phenotyping

Conventional NK cells were isolated from buffy coat using the MACSxpress® Buffy Coat NK Cell Isolation Kit, followed by cryopreservation in CryoStor CS10. On the day of the experiment, frozen cNK and W-NK1 were thawed, washed in medium containing RPMI1640 GlutaMAX™ HEPES + 10% heat inactivated human AB serum (conventional media, CM) and counted. Cells were either immediately (baseline, t = 0h) labelled with receptor binding domains (RBD) derived from envelope glycoproteins of retroviruses (METAFORA Biosystems, Paris, France)[3] or cultured at a density of $1\times10^6$/mL in a 24-well plate for 4h to assess the expression of cell surface nutrient transporters for glucose, lactate, neutral amino acids, cationic amino acids, phosphate, vitamins, riboflavin, and heme. Following culture (see 'Cell culture' paragraph), cells were washed in CM and seeded at 50,000-100,000 cells/well in PBS/1 mM EDTA/5% FBS (FACS buffer). Human TruStain FcX™ was used to block Fc receptors, followed by labeling with RBDs in FACS buffer at 37°C for 20 minutes. Cells were then washed with phosphate buffered saline (PBS) and labelled with CD56, CD16, NKG2A, CD69, CD71 (Miltenyi Biotec, Germany), DAPI (Sigma), along with secondary antibodies (goat anti-mouse IgG1 PE and goat anti-rabbit IgG1 Alexa fluor 647; SouthernBiotech) directed against mouse or rabbit IgG1-Fc-tagged RBDs in FACS buffer at 4°C for 20 minutes. Cells were washed twice with PBS and run through a ZE5 Cell Analyzer (4 lasers with 405, 488, 561 and 640nm excitation; Bio-Rad). Analysis was performed using FlowJo software (BD Biosciences). Live $CD56^+$ cells were gated for the analysis of surface nutrient transporters. Fluorescence minus one (FMO) controls were used to establish background levels in RBD channels (FITC, PE, Alexa Fluor 647).[3]

### Proteomics

Protein extracts from cell lysates or secretome samples (50 µg) were processed through S-trap micro columns (Protifi) according to manufacturer instructions, which include reduction, alkylation and trypsin digestion on-column. Eluted peptides were vacuum-concentrated to dryness and reconstituted in 300 µL at a final concentration of 5% acetonitrile:0.1% formic acid (3 µL, ~ 500 ng protein digest/peptides) and were directly injected by autosampler (Waters M-class) and gradient elution at 10 µL/min (Phenomenex Kinetex XB-$C_{18}$ 2.6 µm 15 x 0.3 mm analytical column, 30°C, mobile phase A 0.1% formic acid:mobile phase B acetonitrile with 0.1% formic acid LCMS grade) onto a Sciex ZenoTOF 7600. Gradient elution with the following profile: 3% B to 35% over 12 min, 80% B at 13-15 min, and re-equilibration at 12 µL/min to the starting conditions at 15.5 min for a 16.5 min total runtime. MS analysis was via the Optiflow source with a micro flow 1-50 µL electrode in positive SWATH mode with zeno pulsing enabled at 4500 V, using 65 variable SWATH windows optimized on a complex human protein lysate from m/z 400-750 12 ms per window following a TOFMS scan of 25 ms for a total cycle time of 1.146 s. Ionization was deactivated at 14.2 min to reduce contamination during the column wash. SWATH data (.wiff format) was processed using DIA-NN 1.8.1 using the FASTA digest for library-free and Deep learning-based spectra options (Swissprot Human May 2022 with addition of TP53 isoforms and the cRAP proteome for contaminants) with the following parameters changed from default: FAST Digest for library-free search and Deep learning on, 1 missed cleavage, max variable mods 1 with Ox(M) selected, heuristic protein inference off. MaxLFQ output from the DIA-NN pg_matrix file[4] was further processed using LIMMA via the AMICA 3.0.1 R based software platform for analysis of proteomics data[5] to generate protein differential expression for downstream analysis, with a min-based imputation.

### Metabolomics

*Cell metabolite extraction*

Cell pellets stored at -80°C were thawed on ice prior to extraction. The cell pellets were washed twice with Phosphate-Buffered Saline (PBS) followed by the addition of 1000μL of ice-cold Methanol (MeOH). Samples were then vortexed and dried. 1000 μL of 2:2:1 MeOH:ACN:$H_2O$ was added to each cell pellet. Samples were then vortexed for 0.5 min and placed in a liquid nitrogen bath for 1 min. The vortex and liquid nitrogen batch steps were repeated three times. Samples were vortexed and placed at -20°C for 1 hr. The following samples were centrifuged at 500 RPM for 10 min at 4°C. Supernatant was then dried. Their protein content was then measured with BCA assay. After the supernatant was dried, 1 μL/2 μg of protein of 2:1 ACN:H2O was added to the residue. Samples were then sonicated at 25°C for 5 minutes and vortexed. The sonication and vortexing was repeated twice. Samples were then stored at 4°C for 1h and centrifuged at 14,000 RPM for 10 min at 4°C. The supernatant was transferred to an LC vial and stored at -80°C until LC/MS analysis.

*LC/MS analysis of polar metabolites*

LC/MS mobile phases A and B were prepared as follows: A) 20 mM ammonium bicarbonate, 0.1% ammonium hydroxide, 5% ACN, 2.5 mM medronic acid and B) 95% ACN. A 4 μL aliquot of polar metabolite extract was analyzed with HILIC/MS by using the following linear gradient at a flow rate of 250 μL/min: 0-1 min: 90% B, 1-12 min: 90-35% B, 12-12.5 min: 35-20% B, 12.5min- 14.5 min: 20% B. The column was re-equilibrated with 20 column volumes of 90% B. Mass spectrometry analysis was completed with a mass range of 67-1500 Da with 1 scan/sec and a mass resolution of 120,000 in both positive and negative ionization mode. MS/MS data was acquired in a data-dependent iterative fashion with a 1.3 m/z isolation window.

*LC/MS analysis of lipid metabolites*

LC/MS mobile phases A and B were prepared as follows: A) 10 mM ammonium formate, 0.1% formic acid, 2.5 mM medronic acid in 6:4 $H_2O$:ACN and B) 10 mM ammonium formate, 0.1% formic acid, 2.5 mM medronic acid in 9:1 IPA:ACN. A 4 μL aliquot of polar metabolite extract was analyzed with HILIC/MS by using the following linear gradient at a flow rate of 250 mL/min: 0-2 min: 30% B, 2-17 min: 30-75% B, 17-20 min: 75-85% B, 20-23 min: 85-100% B, 23-26 min: 100% B, 26-27 min: 100-30% B. The column was re-equilibrated for 5 min. Mass spectrometry analysis was completed with a mass range of 67-1500 Da with 1 scan/sec and a mass resolution of 120,000 in both positive and negative ionization mode.MS/MS data was acquired in a data-dependent iterative fashion with a 1.3 m/z isolation window.

*Metabolite detection and identification*

Metabolite signals (features) were detected in the LC/MS data through in-house peak detection and curation software by Panome Bio (St. Louis, MO). Features were aligned across samples and features with intensities greater than 1/3 of the corresponding intensity in the QC sample were classified as contaminates and removed from future analysis. Feature degeneracy (isotopes, adducts, fragments, etc.) were identified through clustering and ion assignment with in-house software.fMetabolites were structurally identified through searching against a database composed of known metabolites found in RefMet, LipidMaps, and HMDB and comparing isotope patterns and MS/MS fragmentation data (when available). Metabolite identifications were classified based on the Metabolomics Standards Initiative scoring scheme with values ranging from level 1 to level 4. Level 1 identifications are supported by experimental retention times and MS/MS spectra from authentic standards and an isotope pattern match. Level 2 identifications were made on based on isotope pattern matches of greater than 90% similarity (reverse dot-product) and an MS/MS match of greater than 50% similarity (entropy similarity) along with a predicted retention time match of <2 min. Level 3 identifications were made based on an isotope pattern and predicted retention time match with the same cutoffs as defined above. Level 4 identifications are signals determined to correspond to a unique metabolite that did not have any match to a compound in our metabolite database. For lipid compounds, the best matching lipid species is reported. However, the location of double bonds within acyl chains of a lipid species is not discernable from the LC/MS methods utilized. Thus, the double bond positions listed are not meaningful.

*Data normalization and curation*

Metabolomics data from all assays were concatenated. Metabolite signals were discarded if the coefficient of variation (CV) amongst the quality control samples was greater than 25%. Missing values were imputed by using half of the minimum detected intensity for each metabolite. Metabolite intensities were log2 transformed prior to statistical analysis. Metabolite identifications and intensities were manually reviewed for concordance and accuracy. Metabolite identifications and intensities were manually reviewed for concordance and accuracy. Hypothesis testing was executed utilizing a one-way ANOVA, which does not assume equal variances among the groups. Log2 transformed metabolite intensities were used for null hypothesis testing. Fold-changes were computed from non-log2 transformed intensities.

Over-representation analysis was performed with a Fisher's Exact test that compares the expected number of metabolites found to be statistically significant in each pathway with the number of significant metabolites found in each pathway. For null hypothesis testing and pathway analysis, a *P* value cutoff of 0.05 was used.

### Reference mapping of sc-RNA-seq data

ProjecTILs was utilized to accurately project the query dataset (W-NK1 and cNK) onto reference maps constructed from single-cell atlases of annotated NK states using the *make.reference* function with default parameters.[6,7] The *Run.ProjecTILs* function brings the batch-corrected expression matrix of the query dataset into reduced-dimensionality representations of the reference using the anchor-finding and integration algorithms implemented in STACAS and Seurat, which are especially suited to integrate datasets with a potentially large cell type imbalance.[8] After projection, the *cellstate.predict* function was utilized as a nearest-neighbor classifier predicting the state of each query cell by a majority vote of its annotated nearest neighbors in the reference map. Finally, the *plot.statepred.composition* function was employed to visualize the frequency of cell states in the query object as a barplot.

### In vitro functional assays

In vitro cytotoxicity was evaluated against AML cell lines; 10,000 AML cells per well plated on poly-L-ornithine coated 96-well plates, utilizing THP-1-NucLight Red, HL-60-NucLight Red, and TF-1-NucLIght Red as target AML cell lines. Effector cells were plated at E:T ratios of 18:1, 6:1, or 2:1. Additionally, 100 IU/mL of human interleukin-2 (IL-2) was added to the wells, and co-cultures were maintained for 72h at 37°C in a humidified incubator with 5% carbon dioxide ($CO_2$). Live cell imaging was conducted using the IncuCyte S3 Live-Cell Analysis System, capturing images every 3h. Target cell counts were quantified at each timepoint using the IncuCyte S3 analysis software as a measure of W-NK1's cytotoxic activity.

*Cytotoxicity in tumor microenvironment models*

cNK or W-NK1 cells and PKH67-labeled HL-60 cells were mixed at various E:T ratios and resuspended in CM or TME-aligned media. Cell suspension was plated in 96 wells cell plates for 2D condition or embedded in a BME matrix for 3D condition. Cells were incubated for 4h, 24h, 48h or 72h. Cells were recovered from culture wells at the end of each time point and stained for flow cytometry readout. Cells cultured in 2D condition were washed with buffer and stained with fluorescent antibodies cocktail. 3D cultured wells were incubated with collagenase i.v. for 30 minutes for matrix digestion, before being washed and stained with fluorescent antibody cocktails. Responses in terms of tumor killing activity to increasing E:T ratios were fitted using a 4-parameter logistic model equation. Absolute number of cells was normalized with respect to each plate median value of target (no effector) control wells as well as with respect to the number of tumor cells originally seeded (t=0).

*Cytotoxicity in malignant ascites*

HL-60 cells target cells expressing Nuclight Red (Sartorius) were resuspended in malignant ascites or target cell media (DMEM + 20% FBS, Gibco) and seeded into 96-well flat bottom plates (TPP) coated with Poly-L-ornithine (Sartorius). Cryopreserved effector cells were thawed and washed in NK MACS media (Miltenyi Biotec) before being resuspended in malignant ascites or target cell media and plated at the indicated E:T ratios. IL-2 (Miltenyi Biotec) was then added to a final concentration of 100 IU/ml. Target cell survival was assessed in real-time using an IncuCyte Live Cell Analysis System (Sartorius). Live cell numbers were quantified with IncuCyte S3 software and normalized to the number of live cells remaining in the respective target cell-only control group.

*NK cell cytotoxicity under hypoxic conditions*

cNK cells were obtained from a leukapheresis product and isolated using the EasySep™ Human NK Cell Isolation Kit (#17955, StemCell) or W-NK1cells were co-incubated with K562 cells (#CCL-243, ATCC) tagged with CellTrace Violet (ThermoFisher, #C34571). Experiments were performed in an Avatar Incubator (Xcell Biosciences). Briefly, cNK or W-NK1 (effectors) were plated on a 96-well plate at a $1\times10^6$ cells/mL density in complete NK culture media (NK MACS Medium supplemented with 5% AB serum) with or without 500 IU hIL-2/mL and cultured for 24h in a normoxic (5% $CO_2$, atmospheric $O_2$) or hypoxic (5% $CO_2$, 1% $O_2$) condition. Effectors were mixed in wells containing 40,000 K562 (target) cells to achieve final effector-to-target (E:T) ratios of 8:1, 4:1, 2:1 and 1:1. After 24h of incubation, cells were collected and analyzed for cytotoxicity and phenotype using flow cytometry.

### Seahorse real-time cell metabolic analysis

All products for this assay were from Agilent (Cheadle, UK), unless otherwise stated. The Seahorse sensor cartridge was hydrated at 37°C in a non-$CO_2$ incubator overnight, following the manufacturer's instructions. On the day of the assay, cNK cells and W-NK1 were spun onto a Poly D-Lysine (50 µg/mL)-coated XFe24 cell culture microplate with a density of $1\times10^6$ cells per well and rested in Seahorse XF RMPI 1640 medium supplemented with 2 mM L-glutamine, 1 mM sodium pyruvate and 10 mM glucose for 1h in a non-$CO_2$ incubator at 37°C. Extracellular acidification rate (ECAR) and oxygen consumption rate (OCR) were measured using a Seahorse XFe24 extracellular flux analyzer (Agilent Seahorse T Cell Metabolic Profiling Kit). Oligomycin (Thermo 75351; 1 µM) was used to inhibit ATP synthesis; the mitochondrial uncoupler BAM15 (TOCRIS 18756019; 2.5 μM) was injected to produce maximal mitochondrial respiration; and rotenone (Rot; Sigma 557368; 0.5 μM) together with antimycin A (AA; Sigma A8674; 0.5 μM) were injected to derive non-mitochondrial respiration. All samples were run in four replicates.

### In vivo assays

*Trafficking of W-NK1*

Female NSG mice (Jackson Laboratories, MA) were allowed to acclimate on Alfalfa Free food starting 7 days before treatment initiation to minimize background signal (Institutional Animal Care and Use Committee protocol number: EB17-010-063). W-NK1 cells were labelled with IVISense DiR 750 Fluorescent Cell Labeling Dye (Perkin Elmer, Waltham, MA) according to manufacturer's instructions to enable real-time tracking and biodistribution. Mice were dosed *i.v.* with $1\times10^7$ labelled W-NK1 cells resuspended in 100 μL of PBS and biodistribution to peripheral organs was determined by imaging with an AMI HTX at 24h post injection. W-NK1 engraftment and persistence was supported by IL-15 (10 ng/mouse) and IL-2 (50,000 IU/mouse) given i.p. on the day of injection.

*In vivo cytotoxicity against AML cells*

Female NSG (NOD.Cg-Prkdcscid Il2rgtm1Wjl/SzJ) mice (6 – 7 weeks) were purchased from the Jackson Laboratory and maintained in a ventilated caging system with freely available food and sterile water. THP-1 cells (ATCC® Manassas, VA) engineered to express green fluorescent protein (GFP) and click beetle reticulase (CBR) to enable visualization by luminescence were injected i.v. at $0.2\times10^6$ cells/mouse. Successful THP-1 engraftment was confirmed on day four post THP-1 injection by whole-body bioluminescent imaging (BLI). Mice were divided into treatment groups with equivalent average BLI (N = 5 mice/group). W-NK1 treatment was administered either as a single i.v. bolus or three consecutive i.v. injections at weekly intervals starting on D4 post tumor engraftment. Mice received intraperitoneal injections of IL-2 (50,000 IU/mouse) and IL-15 (10 ng/mouse) to support W-NK1 engraftment and persistence. Tumor progression was monitored twice weekly by whole-body BLI measurements and efficacy was expressed as the relative change in flux between vehicle and treatment groups.

### Baseline genotyping and simultaneous MRD and W-NK1 monitoring by NGS

Somatic biomarkers were identified by a custom targeted NGS panel. The 64 most relevant genes involved in myeloid malignancies were screened using DNA samples collected before NK infusion. NGS libraries were prepared from 30ng of DNA using Ion AmpliSeq Kit (ThermoFisher Scientific) and are sequenced with an average coverage of 1500x in an Ion S5 System platform (ThermoFisher Scientific). Variant calling an annotation was performed using Ion Reporter software (version 5.18.2.0). Mutations were called positive when supported by more than nine mutated reads and VAF above 2%. Potential SNPs and germinal variants were excluded. PK-NK analysis was performed by a two-step NGS test. First, a custom amplicon panel containing 54 phased-SNP in linkage-disequilibrium was applied to DNA from both patient and donor NK. Three informative phased-SNP variants on average were selected as donor NK biomarkers. A patient-specific multiplex-PCR was applied to DNA from blood samples after infusion to monitor leukemic blasts and W-NK1 in a single assay. Then, NGS libraries were prepared and sequenced on the S5 System platform with a target sequencing depth of 500,000X per amplicon. A previously described bioinformatic algorithm was used for tumor and NK quantification, including FASTQ file demultiplexing and error-correction algorithm.[9,10] The limit of detection for all somatic SNV, InDels and germline Phased-SNP biomarkers was experimentally defined by analyzing their basal noise in 3 DNA samples from healthy donors. All selected and computable biomarkers had LOD below 0.01% VAF. VAF were normalized to cell count to calculate cell concentration in the sample.

### Key Resources Table

| Software and Bioconductor / R packages |
| --- |

| | | |
|---|---|---|
| clusterProfiler v4.12.0 | Yu et al. (2012) | https://bioconductor.org/packages/clusterProfiler/ |
| Corrplot v0.92 | Wei et al. (2021) | https://github.com/taiyun/corrplot |
| EnhancedVolcano v1.22.0 | Blighe et al. (2019) | DOI:10.18129/B9.bioc.EnhancedVolcano |
| ggplot2 v3.5.1 | NA | https://ggplot2.tidyverseorg |
| Ggrepel v0.9.5 | NA | https://cran.r-project.org/web/packages/ggrepel/index.html |
| Prism v9.0 | GraphPad | https://www.graphpad.com/scientific-software/prism/ |
| R v4.0.4 | R Core Team | https://cran.r-project.org/bin/macosx/ |
| UCell v2.8.0 | Andreatta et al. (2021) | https://github.com/carmonalab/UCell |
| SingleR v2.6.0 | Aran et al. (2019) | https://github.com/LTLA/SingleR |
| Seurat v5.1.0 | Butler et al. (2024) | https://github.com/satijalab/seurat |
| Scran v1.32.0 | Lun et al. (2016) | https://bioconductor.org/packages/scran/ |
| SingleCellExperiment v1.26.0 | Amezquita et al. (2020) | https://bioconductor.org/packages/SingleCellExperiment/ |
| DoubletFinder v2.0.4 | Christopher et al. (2019) | https://github.com/chris-mcginnis-ucsf/DoubletFinder?tab=readme-ov-file |
| Clustree v0.5.1 | Zappia et al. (2018) | https://github.com/lazappi/clustree |
| Scater v1.32.0 | McCarthy et al. (2017) | https://bioconductor.org/packages/scater/ |
| AnnotationDbi v1.66.0 | Pages H et al. (2024) | https://bioconductor.org/packages/AnnotationDbi |
| Limma v3.60.0 | Ritchie ME et al. (2015) | https://bioinf.wehi.edu.au/limma/ |
| MatrixGenerics v1.16.0 | Ahlmann-Eltze C et al. (2024) | https://bioconductor.org/packages/MatrixGenerics |
| Stringr v1.5.1 | Wickham H (2023) | https://github.com/tidyverse/stringr |
| Speckle v0.0.3 | Phipson B et al. (2022) | https://bioconductor.org/packages/speckle/ |
| Scuttle v1.14.0 | McCarthy DJ et al. (2017) | https://bioconductor.org/packages/scuttle/ |
| GenomicRanges v 1.56.0 | Lawrence M et al. (2013) | https://bioconductor.org/packages/GenomicRanges |
| matrixStats v1.3.0 | Henrik B et al. (2024) | https://github.com/HenrikBengtsson/matrixStats |
| org.Hs.eg.db 3.19.1 | Carlson M. (2019) | https://bioconductor.org/packages/org.Hs.eg.db/ |
| S4Vectors 0.42.0 | Pages et al. (2024) | https://bioconductor.org/packages/S4Vectors |

| | | |
|---|---|---|
| miQC v1.12.0 | Hippen et al. (2021) | https://github.com/greenelab/miQC |
| glmGamPoi v1.16.0 | Ahlmann-Eltze et al. (2020) | https://github.com/const-ae/glmGamPoi |
| SummarizedExperiment v1.34.0 | Morgan et al. (2024) | https://bioconductor.org/packages/SummarizedExperiment |
| Tidyverse v2.0.0 | Wickham H | https://github.com/tidyverse/tidyverse |
| GenomeInfoDb v1.40.1 | Arora et al. (2024) | https://bioconductor.org/packages/GenomeInfoDb |
| IRanges v2.38.0 | Lawrence et al. (2013) | https://bioconductor.org/packages/IRanges |
